# Implementation of high-dose glucocorticoids and opioid consumption in the first 24 hours after elective hip and knee arthroplasty: a natural experiment study of 47,317 procedures in Eastern Denmark

**DOI:** 10.64898/2026.08.31.26361793

**Authors:** Jens Laigaard, Marcus Ø Møller, Markus H Olsen, Søren Overgaard, Ole Mathiesen, Anders PH Karlsen

## Abstract

**Background:** In Denmark, high-dose perioperative glucocorticoids were implemented stepwise for patients undergoing total hip arthroplasty (THA), total knee arthroplasty (TKA), and unicompartmental knee arthroplasty (UKA). We aimed to estimate the opioid-sparing effect of adding a single high dose of glucocorticoids to the analgesic regimen.

**Methods:** In this prespecified analysis of a multicentre natural experiment using electronic health record data, we included all elective THA, TKA, or UKA procedures performed in Eastern Denmark from 2017-2025, with no exclusion criteria. At each centre, procedures before implementation of high-dose glucocorticoids served as controls, whereas procedures after implementation comprised the intervention group. A high dose was defined as ≥16 mg or 0.2 mg/kg dexamethasone or ≥80 mg methylprednisolone. The primary outcome was the adjusted between-group difference in 0-24h opioid consumption. The minimal important difference was 5 mg IV morphine equivalents. Secondary outcomes were maximum 0-10 numerical rating scale (NRS) pain score, incidence of opioid-related adverse events, hospital length of stay, and days alive and out of hospital at 30 days.

**Results:** We analysed 47,317 procedures performed at nine centres: 13,010 controls and 34,307 in the intervention group. During the study period, five of nine centres implemented high-dose glucocorticoids for THA patients, two of eight for TKA/UKA patients. High-dose glucocorticoids were administered to 6% and 92% of patients before and after implementation, respectively. The implementation was associated with an adjusted mean reduction of 3.8 mg IV morphine equivalents (95% CI 3.3–4.3) and a 0.8-point lower maximum recorded pain score (99% CI 0.7–0.9) in the first 24 hours, with no statistically significant differences in the remaining outcomes.

**Conclusions:** Implementation of high-dose glucocorticoids was associated with only a small drop in opioid consumption, below the minimal important difference.

**Key Points Summary:**

- **Question**: What opioid-sparing effects may be expected when adding high-dose glucocorticoids (≥16 mg or 0.2 mg/kg dexamethasone or ≥80 mg methylprednisolone) to the standard analgesic package for patients undergoing total hip and knee arthroplasty?
- **Findings**: In Eastern Denmark, the adoption of high-dose glucocorticoids to the existing multimodal analgesic regimen was associated with a slightly lower 0-to-24 hours in-hospital opioid consumption, according to electronic health record data.
- **Meaning**: Clinicians may expect a slightly lower opioid-sparing effect from high-dose glucocorticoids than found in randomised clinical trials.

## Introduction

Although there is ongoing debate about the optimal dosing, most guidelines recommend that glucocorticoids are administered to patients undergoing total hip arthroplasty (THA), total knee arthroplasty (TKA), and unicompartmental knee arthroplasty (UKA).^1–3^ The aim is to prevent nausea, reduce inflammation and pain, and enhance early recovery. This recommendation is backed by evidence from large randomised clinical trials (RCTs) and subsequent meta-analyses; however, there is relatively sparse evidence from ‘real-world’ studies.^4–6^

While RCTs continue to be the most important design in comparative research, there is a growing appreciation of analyses based on clinical data.^7^ Randomisation, blinding, and intention-to-treat (ITT) principles ensure unbiased estimates of the effect of assigning the trial participants to the intervention group. However, even pragmatic RCTs exclude a substantial proportion of patients that would be treated in clinical practice, such as patients with chronic opioid use.^5,8,9^ Also, participants in RCTs are monitored more closely and may be treated differently than in the clinical setting, e.g. opioids administered through patient-controlled analgesia instead of nurse-administered.^8,10^ These differences may influence the observed effects of the intervention.

Since 2011, more and more centres in Denmark have adopted high-dose glucocorticoids, typically methylprednisolone 125 mg or dexamethasone 24 mg, as part of the routine analgesic package for patients undergoing THA, TKA, and UKA.^11,12^ This has generated a series of natural before-and-after experiments, which enabled us to evaluate the adoption of the high-dose glucocorticoids using data from the electronic health records.^13^ In this natural experiment study, we aimed to examine the opioid-sparing effect of adding high-dose glucocorticoids to the existing analgesic regimen for patients undergoing primary THA, TKA, and UKA.

## Methods

This was a predefined analysis of a natural experiment using routinely collected data from the electronic health record. The study was reported following the Strengthening the Reporting of Observational Studies in Epidemiology (STROBE) guideline.^14^ This study was listed at the Capital region of Denmark’s regional research listing (p-2025-20115; October 28, 2025), the Ethical Committee of the Capital Region of Denmark waived ethical approval for this study (identifier F-25000312, January 2, 2025), and we were granted access to individual patient files from the Capital Region Team for Medical Records Data (R-25000339, August 21, 2025).

### Study setting

The patients considered for this study underwent primary THA, TKA, and UKA at public centers in the Capital Region and Region Zealand in Denmark from 2017 to 2025. The centers were the orthopedic departments at the following hospitals: Bispebjerg Hospital, Bornholms Hospital, Gentofte Hospital, Hvidovre Hospital, North Zealand Hospital, Holbæk Hospital, Central and West Zealand Hospital Næstved, Zealand University Hospital Køge, and Zealand University Hospital Nykøbing Falster.

### Eligibility criteria

The unit of analysis was procedures, and all primary, elective THA, TKA, and UKAs performed in adults (>18 years) from 2017 to 2025 were eligible. There were no exclusion criteria.

### Intervention

The intervention was the addition of high-dose glucocorticoids to the standard analgesic package. A high-dose steroid dose was defined as intravenous (IV)/oral administration of at least 16 mg or 0.2 mg/kg dexamethasone or at least 80 mg methylprednisolone. The intervention was given along with other routine analgesic interventions. The comparator was lower or no doses of glucocorticoids.

### Assignment of intervention

Treatment allocation was based on whether the centre where the patient underwent surgery had implemented the intervention at the time of surgery. For each centre, procedures performed *before* implementation of glucocorticoids served as controls, while procedures *after* implementation constituted the intervention group, regardless of their actual treatment. The date of implementation was determined from plots of the proportion of patients receiving high-dose glucocorticoids over time as the point that visually best separated the two groups.

### Outcomes

The primary endpoint was the between-group difference in opioid consumption in the first 24 hours after surgery, expressed in IV morphine milligram equivalent doses. Apart from remifentanil, opioid doses administered pre-emptively at the end of surgery were included in the outcome, adjusting for time interval between administration and the actual end of surgery using the drug-specific half-life.^15^ We used the opioid conversion table that is implemented in the TRIPLE-A platform for conversions to morphine equivalents.^16^ The minimal important difference was set at 5 mg IV morphine equivalents.^17^

Secondary endpoints: a) Worst postoperative pain intensity during 0-24 hours after surgery, measured with the 0-10 numerical rating scale (NRS); b) Proportion of patients with at least one opioid-related adverse event during 0-24 hours after surgery; c) Length of stay in the post-anaesthesia care unit, measured in minutes; d) Hospital length of stay, measured in hours from end of surgery to hospital discharge; and e) Days alive and out of hospital at 30 days.

### Data sources

We used data available from the AI and Automation in Anaesthesia (TRIPLE-A) database.^15^ The database includes hundreds of variables on well over one million surgical procedures performed at public centres in the Capital Region and Region Zealand in Denmark from 2017 to 2025, extracted from the electronic health record system Sundhedsplatformen (Epic Systems Corporation, WI, USA).

### Statistical Analysis

We followed the predefined statistical analysis plan (https://doi.org/10.1101/2025.11.11.25339982, last updated April 18, 2026) mimicking the statistical methods used in stepped-wedge cluster-RCTs. All analyses were conducted in R (Version 4.2.2, R Core Team, Vienna, Austria). The key assumption was that, at the time of implementation of high-dose glucocorticoids into the standard analgesic package, no other changes in patient selection, treatment, or outcome assessment occurred at that centre.^13^ This assumption was checked with statistical process control, described below.

#### Sample size

A priori, we used the CRTsize package to calculate an estimate of power. We anticipated a power of 95% (5% false-negative rate) for detection of a 5 mg IV morphine equivalents difference, assuming a standard deviation (SD) of 10, 14 paired clusters (i.e. 7 centers) with 50 patients, an intraclass coefficient of 0.1, and a 5% false-positive rate.

#### Missing data

There was substantial missing outcome data for worst postoperative pain intensity during 0-24 hours after surgery, with 28% missing in the control group and 27% missing in the intervention group. Because the missingness was balanced, we performed multiple imputation by chained equations with predictive mean matching. Similarly, 45 missing procedures had missing opioid-related adverse event data (control group: 0.0%, intervention group: 0.1%), which were also imputed. We also had missing outcome data for length of stay in the post-anaesthesia care unit, with 0.5% missing in the control group and 8.5% missing in the intervention group. As specified in the statistical analysis plan, this outcome was not analysed because the missingness was severely unbalanced between groups.

After accessing the data, we decided to exclude extreme outliers in terms of intraoperative opioid use (≥130 mg IV morphine equivalents), and opioid consumption in the first 24 hours after surgery (≥300 mg IV morphine equivalents) because of a strong suspicion of erroneous data registration.

#### Modelling

The main analyses was based on a pre-defined mixed-effects model using the lme4 package, which included a before-after variable, indicating if high-dose glucocorticoids were implemented at the at time of surgery, a categorical time variable with a level for each interval between time of implementation, age, sex, type of surgery (THA/TKA/UKA), type of anesthesia (general/spinal/general and nerve block), count of non-opioid analgesics (acetaminophen/NSAID/gabapentin), local infiltration analgesia, preoperative chronic opioid use, psychiatric diagnosis, and centre (random intercept).^18^ Although we had planned to use the *count* of psychiatric diagnoses as a covariable, the prevalence of multiple registered psychiatric diagnoses was low, so the variable was simplified to a binary variable.

#### Thresholds for significance

The main conclusion was based on the primary analysis of the primary outcome, i.e. in the ITT population, at the 0.05 significance level. Continuous outcomes were reported as adjusted mean difference with 95% (primary outcome) or 99% (secondary outcomes) confidence intervals (CIs). Binary outcomes (i.e. opioid-related adverse events) were reported as adjusted absolute risk differences with 99% confidence intervals.

#### Sensitivity analyses

We also conducted per-protocol sensitivity analyses of all outcomes, excluding patients in the control group if they received the high-dose glucocorticoids and patients in the intervention group if they failed to receive high-dose glucocorticoids.

We conducted several post-hoc sensitivity analyses, to assess the robustness of our findings. First, the THA and TKA/UKA cohorts were also analysed separately, resetting the categorical time variable to only have one level per interval between implementation dates within that cohort.

Second, our main model includes a substantial number of covariables to control for unforeseen bias, putting it at risk of overfitting. We therefore conducted a less adjusted analysis, adjusting only for time, type of surgery, and centre.^13^ Third, we imputed data for two outcomes, pain score and opioid-related adverse events, and therefore conducted complete-case analyses of these outcomes. Fourth, count of non-opioid analgesics was included as a continuous count variable. Because this approach assumes that the effect of acetaminophen, NSAID and gabapentin is identical, we also conducted an analysis including them as individual binary variables. Fifth, we observed a substantial difference in the number of patients discharged within 24 hours after surgery between the groups. Because our data was restricted to in-hospital opioid consumption, we therefore conducted sensitivity analyses excluding patients discharged within 24 hours.

#### Statistical process control charts

We used statistical process control in control charts with monthly data relative to the time of implementation to visualise the findings and underlying trends in patient characteristics, co-interventions, and outcomes. We applied the Anhøj rules for *unusually* long runs and *unusually* few crossings of the median and the more traditional Shewhart’s 3-sigma rule (values outside +/- 3 SDs) to assess for non-random patterns in the distribution of data points.^19^

#### Data sharing

No data were collected for this specific study. The full individual participant data used in this project are available from the TRIPLE-A database with a signed data transfer/access agreement and approval from the Capital Region Team for Medical Records Data.^15^ The full statistical code can be obtained from the corresponding author upon request.

## Results

We included 47,390 primary, elective THA, TKA, and UKA procedures performed in 40,772 adults. Of these, 57 (0.2%) and 16 (0.1%) were excluded as outliers from the intervention and control group, respectively. Thus, we analysed a total of 47,317 procedures performed at nine centres: 34,307 in the intervention group and 13,010 in the control group (**Figure 1**).

**Figure 1:**
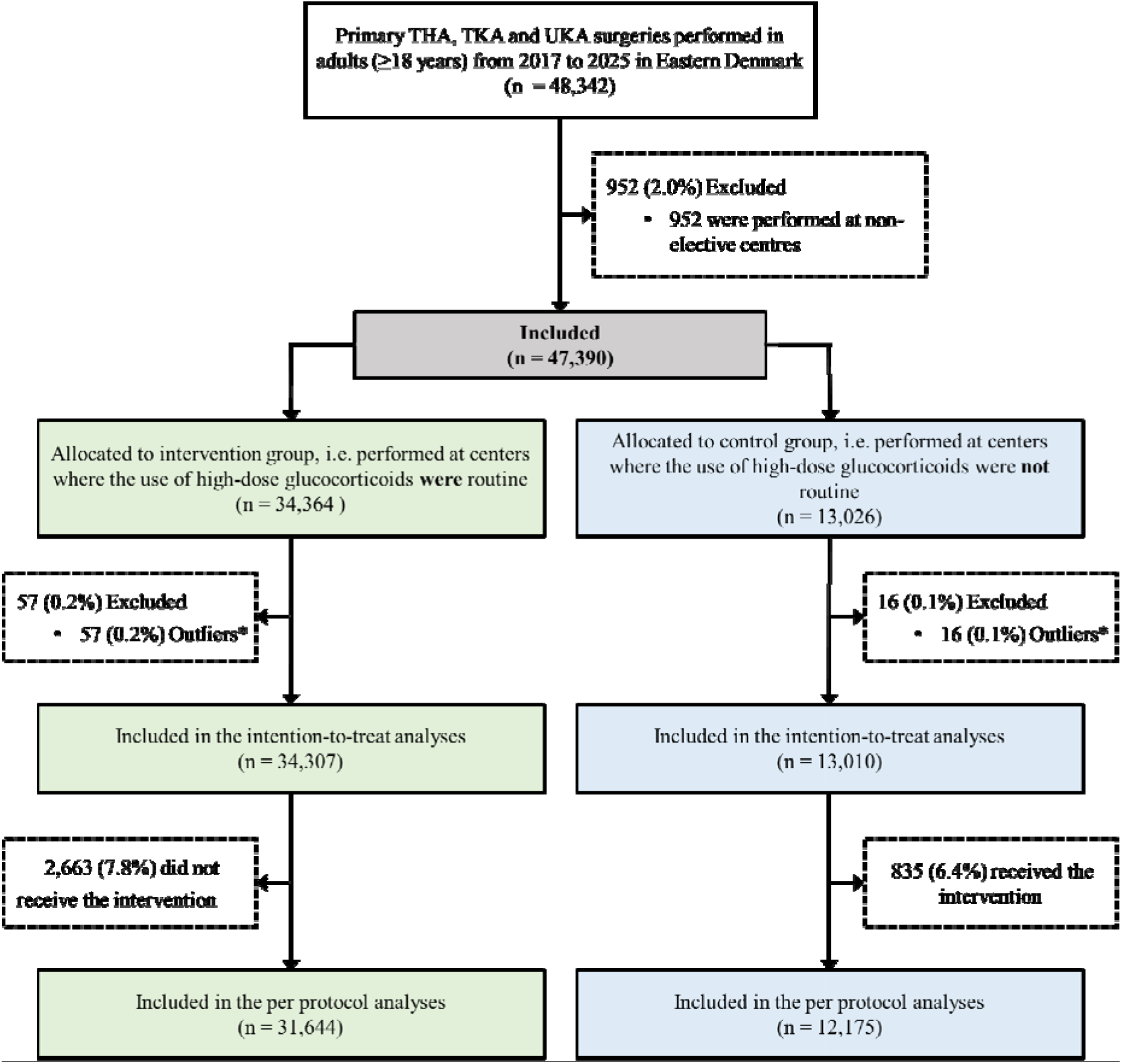
Participant flow diagram. *Outliers were procedures with extreme intraoperative opioid use (≥130 mg intravenous morphine equivalents) or extreme opioid consumption in the first 24 hours after surgery (≥300 mg intravenous morphine equivalents), most likely reflecting erroneous data registration. THA = Total Hip Arthroplasty, TKA = Total Knee Arthroplasty, UKA = Unicompartmental Knee Arthroplasty

During the study period, five of nine centres implemented high-dose glucocorticoids for patients undergoing THA, and two of eight centres implemented high-dose glucocorticoids for patients undergoing TKA and UKA. The remaining centres used high-dose glucocorticoids as part of their standard analgesic package throughout the study period and were included in the analyses as reference cohorts (**Figure 2**).

**Figure 2:**
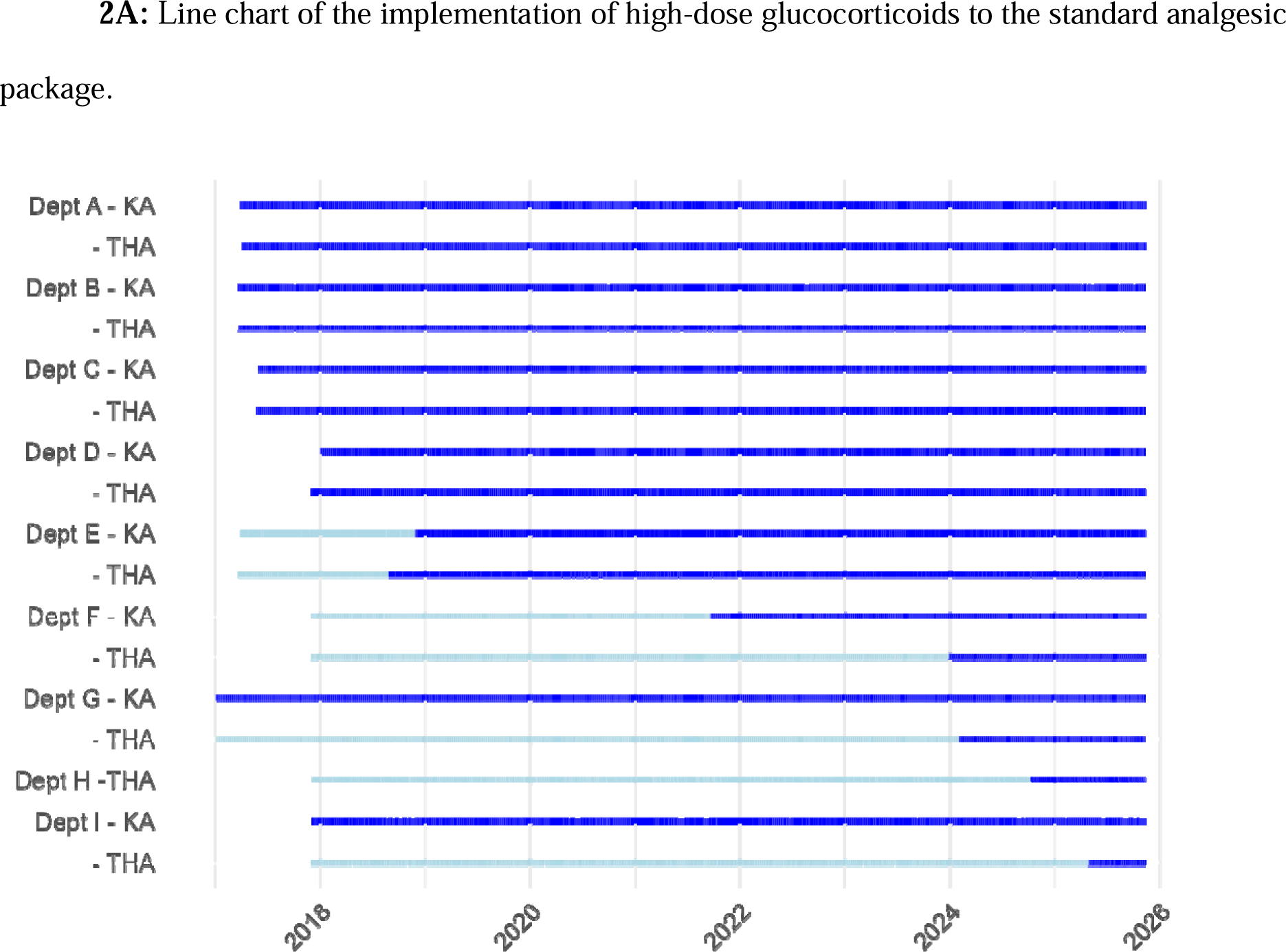
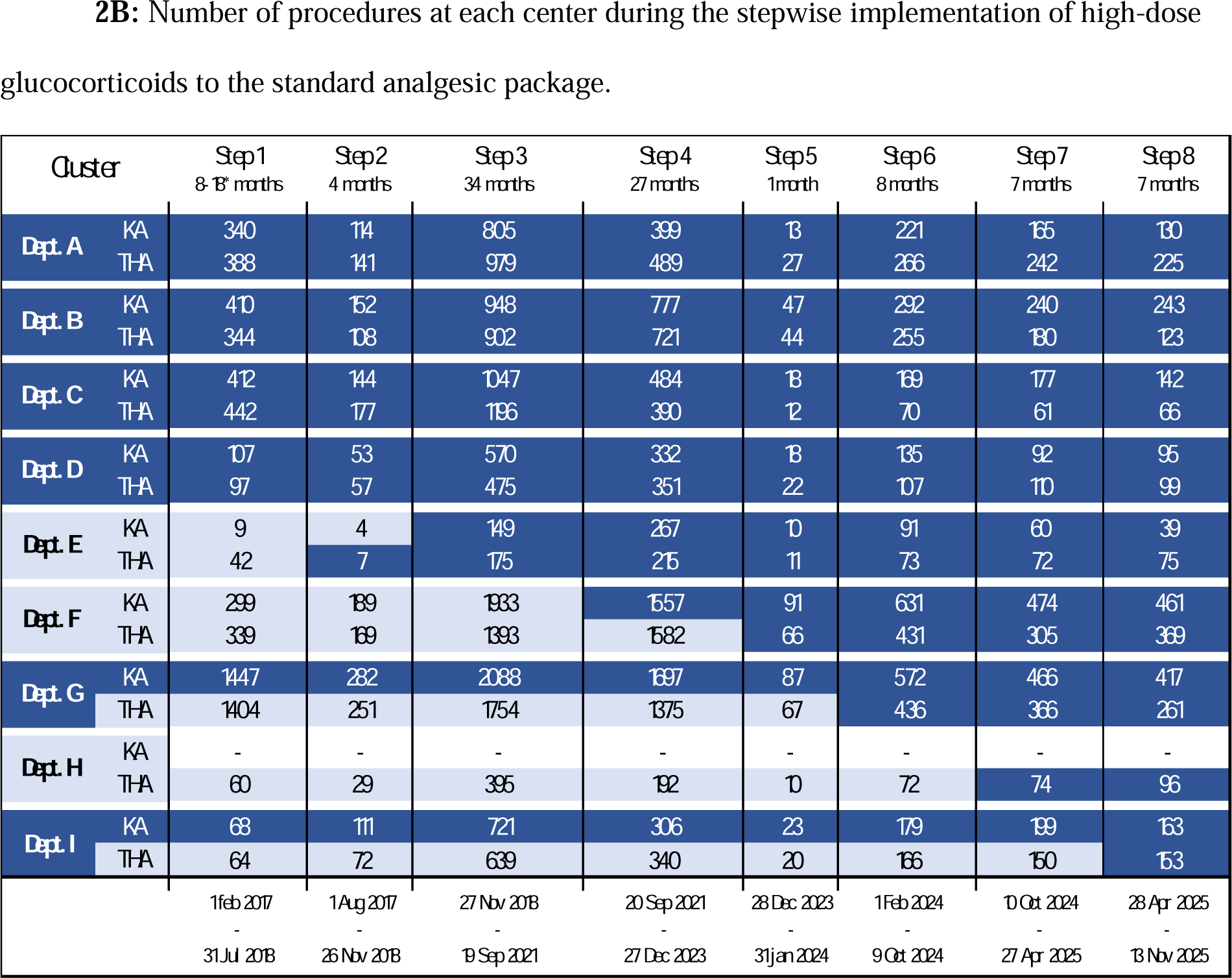
Light blue lines and fields illustrate that high-dose glucocorticoids have not yet been implemented, while dark blue lines and fields illustrate routine use. Department A, B, C, D, G (KA) and I (KA) implemented high-dose glucocorticoids before the study period began but are included in the analyses as reference cohorts to control for underlying trends. *Step 1 varies in length because the centers in Region Zealand adopted the electronic health record system 10 months after the Capitol Region. THA = total hip arthroplasty, KA = knee arthroplasty.

### Participants

The cohort was mostly female (60%) with a mean age of 70 years. Most patients had spinal anaesthesia (74%), and many received local infiltration analgesia (67%). The intervention and control groups had comparable patient characteristics, including age, sex, presurgical opioid use, and the prevalence of a psychiatric diagnosis (**Table 1**). At centres that implemented the intervention, substantially more patients in the intervention group were discharged within 24 hours (41% versus 18% in the control group).

**Table 1.**
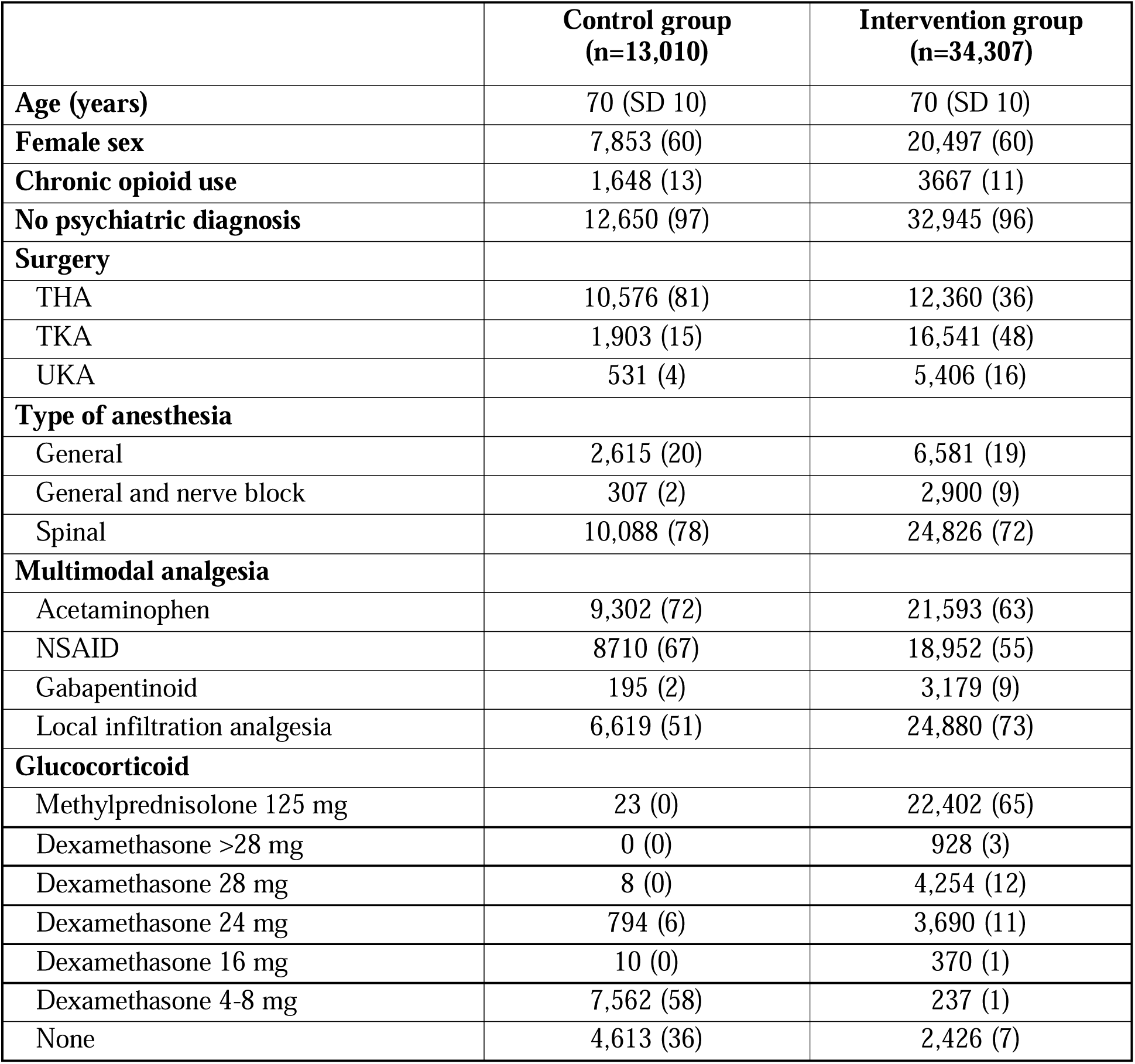
Baseline characteristics. Number of patients (% of all patients). THA = total hip arthroplasty, TKA = total knee arthroplasty, UKA = unicompartmental knee arthroplasty, NSAID = non-steroidal anti-inflammatory drug, mg = milligram.

After adopting the intervention to the routine analgesic package, 92% of patients received high-dose glucocorticoids. In comparison, 6% of patients received high-dose glucocorticoids before its implementation (**Figure S1**).

Co-interventions, including the use of gabapentinoids, nerve blocks, and local infiltration analgesia, were unbalanced, which was expected since different centres contributed different numbers of patients to the two groups. Statistical process control charts showed little variation in patient demographics and co-interventions relative to the implementation of high-dose glucocorticoids.

This finding supports the assumption that the implementation was not accompanied by concurrent changes in patient selection or co-interventions (**Figure S2a-j**). However, in the centres that implemented high-dose glucocorticoids during the study period, premedication with NSAIDs increased from approximately 15 months post-implementation.

The patient characteristics and co-interventions are also presented stratified by type of surgery (**Table S1**) and restricted to centres that implemented the intervention during the study period (**Table S2**).

### Outcomes

The mean opioid consumption in the control group was 22.1 mg IV morphine equivalents (SD 22.3) in the first 24 hours following surgery compared to 20.7 mg IV morphine equivalents (SD 21.2) in the intervention group, with large variation across centres (**Figure 3)**. In the ITT analysis, implementation of high-dose glucocorticoids was associated with an adjusted mean reduction of 3.8 mg IV morphine equivalents (95% CI 3.3 to 4.3) (**Table 2**), which was below the prespecified minimal important difference of 5 mg IV morphine equivalents. The estimate was slightly larger in the per-protocol population (−5.0 mg IV morphine equivalents; 95% CI −4.5 to − 5.5) (**Table 3**), but similar in sensitivity analyses (**Table S3**). The difference was not clearly visible in the statistical process control chart (**Figure S3a**). Post-hoc subgroup analyses of THA and TKA/UKA procedures yielded uncertain results due to fewer shifts in treatment. In both subgroups, the implementation of high-dose glucocorticoids was associated with very small reductions in 0-to-24-hour opioid consumption (**Table S4)**.

**Figure 3:**
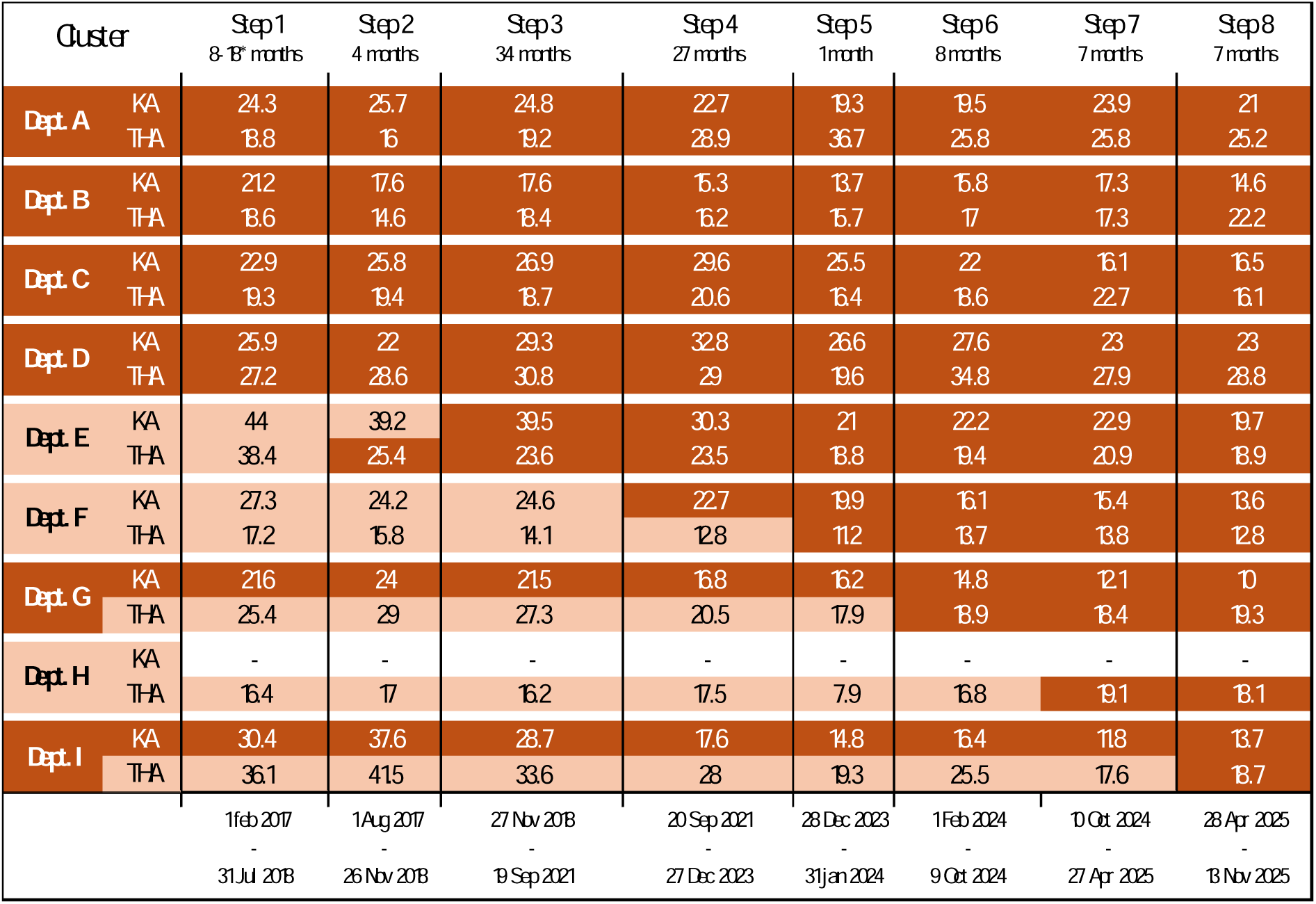
0-to-24-hour opioid consumption at each center during the stepwise implementation of high-dose glucocorticoids to the standard analgesic package. Light orange fields illustrate that high-dose glucocorticoids have not yet been implemented, while dark orange fields illustrate routine use. *Step 1 varies in length because the centers in Region Zealand adopted the electronic health record system 10 months after the Capitol Region. THA = total hip arthroplasty, KA = knee arthroplasty.

**Table 2:**
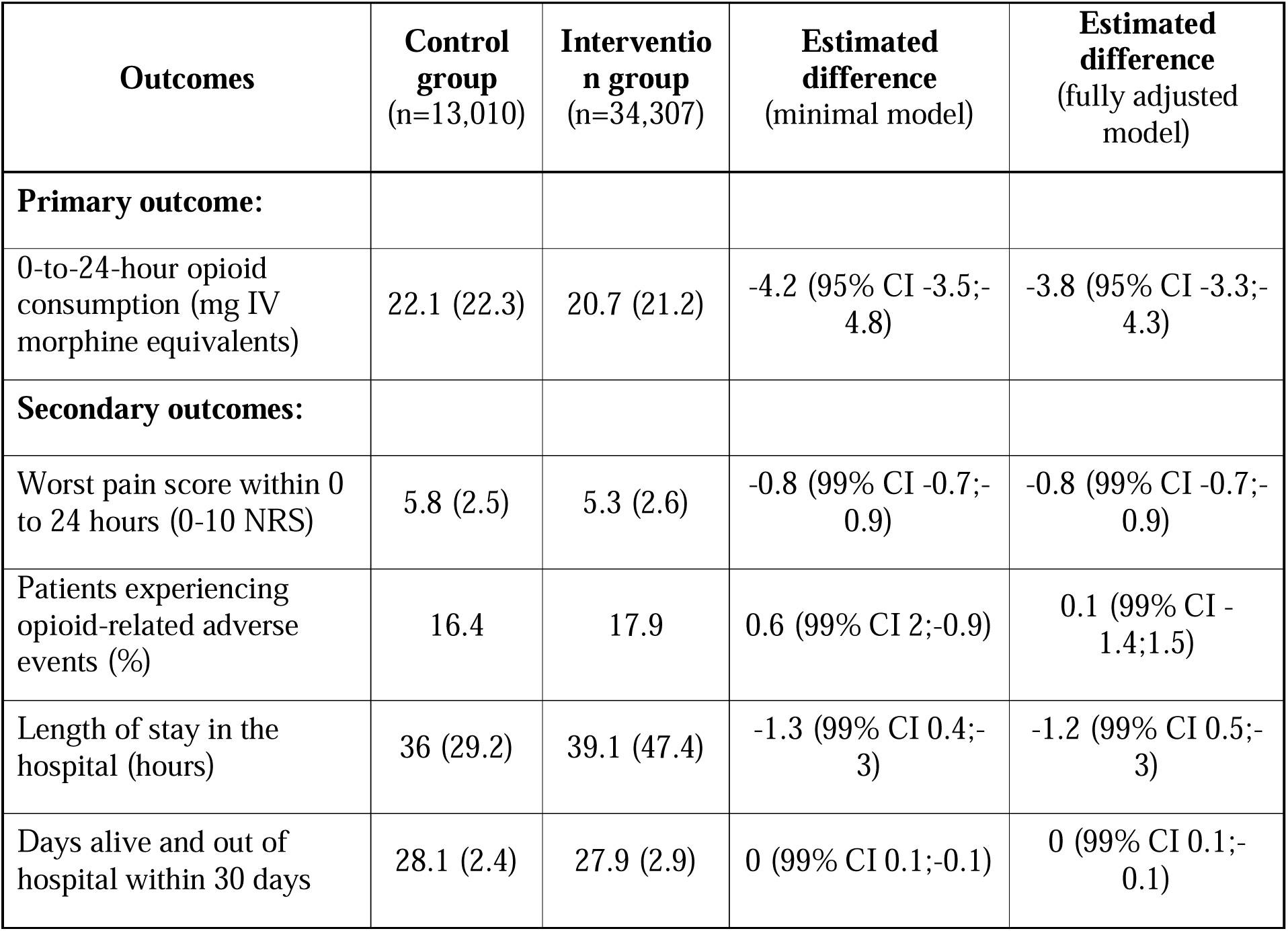
Primary and secondary outcomes in the intention-to-treat population. Values are mean (standard deviation) or incidence. Estimates are average treatment effects. The minimal model is adjusted for time period, type of surgery, and center (random effect). The fully adjusted model is adjusted for age, sex, time period, type of surgery, type of anesthesia, number of other multimodal analgesics, local infiltration analgesia, chronic opioid use, psychiatric diagnoses, and center (random effect). IV = Intravenous, CI = Confidence interval, NRS = Numerical rating scale score

**Table 3:** Primary and secondary outcomes per-protocol population. Values are mean (standard deviation) or incidence. Estimates are average treatment effects. The minimal model is adjusted for time period, type of surgery, and center (random effect). The fully adjusted model is adjusted for age, sex, time period, type of surgery, type of anesthesia, number of other multimodal analgesics, local infiltration analgesia, chronic opioid use, psychiatric diagnoses, and center (random effect). IV = Intravenous, CI = Confidence interval, NRS = Numerical rating scale score

| Outcomes | Control group<br>(n=12,175) | Intervention group<br>(n=31,644) | Estimated difference<br>(minimal model) | Estimated difference<br>(fully adjusted model) |
| --- | --- | --- | --- | --- |
| **Primary outcome:** |  |  |  |  |
| 0-to-24-hour opioid consumption (mg IV morphine equivalents) | 23.1 (22.4) | 20.6 (21.2) | -5.7 (95% CI -5;-6.3) | -5 (95% CI -4.5;-5.5) |
| **Secondary outcomes:** |  |  |  |  |
| Worst pain score within 0 to 24 hours (0-10 NRS) | 5.8 (2.5) | 5.3 (2.6) | -0.9 (99% CI -0.8;-1) | -0.9 (99% CI -0.8;-1) |
| Patients experiencing opioid-related adverse events (%) | 16.6 | 17.7 | -0.1 (99% CI 1.4;-1.6) | -0.6 (99% CI -2.1;0.9) |
| Length of stay in the hospital (hours) | 36.2 (28.8) | 38.5 (44.5) | -2.4 (99% CI -0.7;-4.1) | -2.2 (99% CI -0.4;-3.9) |
| Days alive and out of hospital within 30 days | 28.1 (2.4) | 27.9 (2.8) | 0.1 (99% CI 0.2;0) | 0.1 (99% CI 0.2;-0.1) |

The intervention was associated with a lower maximum pain score during the first 24 hours (−0.8 points; 99% CI −0.7 to −0.9), with similar results in the per-protocol population. In the ITT population, there was no significant difference between groups with respect to the proportion of patients experiencing an opioid-related adverse event, hospital length of stay, or days alive and out of hospital at 30 days. In the per-protocol population, the intervention was associated with a shorter length of stay in the hospital (−2.2 hours; 99% CI −0.4 to −3.9). The statistical process charts showed no clear trends or changes in any of the secondary outcomes (**Figure S3b-e**). For the two outcomes that had missing outcome data, maximum pain score and opioid-related adverse events, the complete case sensitivity analyses gave similar results (**Table S3**).

## Discussion

In this analysis of a natural experiment, we found that the implementation of high-dose glucocorticoids to routine clinical practice was associated with statistically significant but modest reductions of opioid requirements and pain intensity in the first 24 hours after primary THA, TKA, and UKA. Both differences were below the prespecified thresholds for clinical importance, and there was no statistically significant difference in opioid-related adverse events, hospital length of stay, or days alive and out of hospital.

In this study, the adoption of high-dose glucocorticoids to routine practice was associated with slightly lower 0–24-hour opioid-sparing and pain-relieving effects than found in large previous RCTs.^4–6^ This may be explained by several factors. First, we imposed no exclusion criteria, meaning that our population most likely differed from most RCT populations – even pragmatic ones.^6,8^ Second, we assessed opioid use administered by the clinical staff, which mainly includes extended-release opioids and around-the-clock administrations. In contrast, RCT participants often exclusively receive immediate-release opioids through patient-controlled analgesia, making the RCTs more sensitive to change.^20^ Third, a subgroup of patients in the control group received the intervention, many as part of the RECIPE or DEX2TKA trials which ran during the study period, both using dexamethasone 24 mg.^5,6^ Similarly, a substantial proportion of patients in the intervention group did not receive the intervention. Fourth, the intervention was mainly implemented in patients undergoing THA who use significantly fewer opioids than patients undergoing TKA or UKA, which means that it is harder to achieve an absolute opioid-sparing effect of 5 mg IV morphine equivalents.^4,21^ Last, compared to the minimal model, the main model trended towards a smaller difference across all outcomes, which may indicate overfitting of data with the fully adjusted model and therefore an underestimation of the estimate.

Regarding safety, we found no evidence to suggest that a single high dose of glucocorticoids was harmful. The main ITT analyses showed no statistically significant difference in hospital length of stay or days alive and out of hospital at 30 days between groups, while the per-protocol analyses suggested a slightly shorter hospital stay with the intervention. Most hospitals applied enhanced recovery after surgery (ERAS) principles during the study period, and length of stay was therefore generally short, limiting the ability to detect potential differences in this outcome. We acknowledge that relying solely on process outcomes prevents us from drawing conclusions on specific adverse effects such as hyperglycaemia or insomnia. We choose these outcomes because timestamp-based variables are less susceptible to spurious findings caused by changes in registration practices. Although not specifically powered towards safety, previous systematic reviews and large RCTs have not found evidence that glucocorticoid treatment increases the risk of adverse events, including infections, fractures or impaired wound healing.^4–6,22^

The study employed a powerful natural experiment design. The study was multi-centred and employed no exclusion criteria, which means that our results are highly generalisable to settings with high compliance to ERAS guidelines.^12^ The intervention was quite abruptly implemented as it became part of the routine analgesic package for patients undergoing THA, TKA, and UKA. Thus, in contrast to many stepped-wedge studies, we did not require a transition period. The robustness of our findings is supported by the consistency of findings across ITT, per-protocol, and sensitivity analyses. Particularly, adjusting for a range of patient-level risk factors and co-interventions only changed the estimates slightly, which supports the causal inference. Still, the interpretation of the study is limited mainly by its observational design and the use of routinely collected data, which has missingness and may contain errors. Although we attempted to mitigate bias statistically, there were no blinding and we were unable to account for organizational changes, that we may have failed to observe.

The analysis was predominantly based on patients undergoing THA, because more centres implemented high-dose glucocorticoids for these patients during the study period. Thus, our results may be more applicable to the THA population. Similarly, both the implementation of methylprednisolone and dexamethasone was analysed collectively, which limits the results interpretation. Because the intervention was incompletely implemented, the estimates from the ITT analyses reflect the effect of adding high-dose glucocorticoids to the routine multimodal analgesic regimen, averaged across all patients. Since not all patients will receive the intervention, this estimate is lower than the treatment effect that can be expected in individual patients, i.e. the per-protocol effect. Yet the per-protocol estimates in the current study may be confounded by indication, e.g. patients who were not given high-dose glucocorticoids after its implementation may be different from those who were. Last, the certainty of our estimates is limited by the relatively low number of centres that implemented the intervention during the study period, of which some contributed relatively few patients.

### Conclusion

We found that the adoption of high-dose glucocorticoids into routine clinical practice was associated with a 3.8 mg IV morphine equivalents drop in opioid consumption in the first 24 hours after primary THA, TKA, and UKA. This estimate was below the prespecified minimal important difference. The intervention also reduced the maximum pain score by 0.8 points, while there was no statistically significant difference in the incidence of opioid-related adverse events, hospital length of stay, or days alive and out of hospital at 30 days. The analyses depended predominantly on data from patients undergoing THA, and the results may therefore be more applicable to this population.

## Online Registration

https://doi.org/10.1101/2025.11.11.25339982

## Presentations

Abstract submitted for The Nordic Orthopaedic Federation/Danish Orthopaedic Society Congress 2026 in Copenhagen (11-13 Nov 2026) and for The Danish Society of Anaesthesiology and Intensive Care Medicine’s annual meeting (12-14 Nov 2026).

## Data Availability

No data were collected for this specific study. The full individual participant data used in this project are available from the TRIPLE-A database with a signed data transfer/access agreement and approval from the Capital Region Team for Medical Records Data. The full statistical code can be obtained from the corresponding author upon request.

## Acknowledgments

The project was supported by a grant from The Danish Society of Anaesthesiology and Intensive Care Medicine’s Research Initiative. Mistral Vibe (Mistral AI, Paris, France) was used to assist with language editing of individual sentences.

## Funding Statement

This project was supported by a grant from The Danish Society of Anaesthesiology and Intensive Care Medicine. None besides the authors had influence on the design, analysis, or dissemination of the study.

## Conflict of Interest

The authors declare no competing interests.

## Notes

### Competing Interest Statement

The authors have declared no competing interest.

### Clinical Protocols

https://doi.org/10.1101/2025.11.11.25339982

### Author Declarations

The Ethical Committee of the Capital Region of Denmark waived ethical approval for this study (identifier F-25000312, 2 January 2025).

### Summary of Updates

Updated with more appropriate interpretation and wording (i.e. "association" not "cause and effect")

