## Supplementary material for "Implementation of high-dose glucocorticoids and opioid consumption in the first 24 hours after elective hip and knee arthroplasty: a natural experiment study of 47,317 procedures in Eastern Denmark": Online Supplementary Materials

### Table S1: Baseline characteristics of participants stratified by type of surgery

|  | **Knee Arthroplasty** | | **Total Hip Arthroplasty** | |
| --- | --- | --- | --- | --- |
|  | **Control  (n=2,434)** | **Intervention  (n=21,947)** | **Control  (n=10,576)** | **Intervention  (n=12,360)** |
| **Age (years)** | 68 (SD 9) | 69 (SD 10) | 70 (SD 10) | 70 (SD 11) |
| **Female sex** | 1,382 (57) | 12,910 (59) | 6,471 (61) | 7,587 (61) |
| **Chronic opioid use** | 286 (12) | 2,107 (10) | 1,362 (13) | 1,560 (13) |
| **No psychiatric diagnosis** | 2,368 (97) | 21,072 (96) | 10,282 (97) | 11,873 (96) |
| **Surgery** |  |  |  |  |
| TKA | 1,903 (78) | 16,541 (75) | - | - |
| UKA | 531 (22) | 5,406 (25) | - | - |
| **Anaesthesia** |  |  |  |  |
| General | 309 (13) | 3,418 (16) | 2,306 (22) | 3,163 (26) |
| General and nerve block | 91 (4) | 2,569 (12) | 216 (2) | 331 (3) |
| Spinal | 2,034 (84) | 15,960 (73) | 8,054 (76) | 8,866 (72) |
| **Multimodal analgesics** |  |  |  |  |
| Acetaminophen | 1,904 (78) | 14,621 (67) | 7,398 (70) | 6,972 (56) |
| NSAID | 2,269 (93) | 14,659 (67) | 6,441 (61) | 4,293 (35) |
| Gabapentinoid | 48 (2) | 1,529 (7) | 147 (1) | 1,650 (13) |
| Local infiltration analgesia | 1,847 (76) | 18,491 (84) | 4,772 (45) | 6,389 (52) |
| **Glucocorticoid** |  |  |  |  |
| Methylprednisolone 125 mg | 1 (0) | 14755 (67) | 22 (0) | 7647 (62) |
| Dexamethasone >28 mg | 0 (0) | 688 (3) | 0 (0) | 240 (2) |
| Dexamethasone 28 mg | 1 (0) | 3020 (14) | 7 (0) | 1234 (10) |
| Dexamethasone 24 mg | 233 (10) | 1715 (8) | 561 (6) | 1975 (16) |
| Dexamethasone 16 mg | 1 (0) | 113 (0) | 9 (0) | 257 (2) |
| Dexamethasone 4-8 mg | 1852 (76) | 119 (1) | 5710 (54) | 118 (1) |
| None | 346 (14) | 1537 (7) | 4267 (40) | 889 (7) |

Number of patients (% of all patients), except for Age, which is mean (SD). TKA = total knee arthroplasty, UKA = unicompartmental knee arthroplasty, NSAID = non-steroidal anti-inflammatory drug, mg = milligram.

### Table S2: Baseline characteristics of participants who underwent surgery at clusters that implemented high-dose glucocorticoids

|  | Control group (n=13,010) | Intervention group (n=7,024) |
| --- | --- | --- |
| **Age (years)** | 70 (SD 10) | 70 (SD 10) |
| **Female sex** | 7,853 (60) | 4,114 (59) |
| **Chronic opioid use** | 1,648 (13) | 462 (7) |
| **No psychiatric diagnosis** | 12,650 (97) | 6,786 (96) |
| **Surgery** |  |  |
| Total hip arthroplasty | 10,576 (81) | 3,194 (46) |
| Total knee arthroplasty | 1,903 (15) | 2,753 (39) |
| Unicompartmental knee arthroplasty | 531 (4) | 1,077 (15) |
| **Type of anaesthesia** |  |  |
| General | 2,615 (20) | 1,641 (23) |
| General and nerve block | 307 (2) | 165 (2.3) |
| Spinal | 10,088 (78) | 5,218 (74) |
| **Multimodal analgesics** |  |  |
| Acetaminophen | 9,302 (72) | 5,363 (76) |
| NSAID | 8,710 (67) | 5,513 (79) |
| Gabapentinoid | 195 (2) | 188 (3) |
| Local infiltration analgesia | 6,619 (51) | 4,685 (67) |
| **Glucocorticoid** |  |  |
| Methylprednisolone 125 mg | 23 (0) | 2044 (29) |
| Dexamethasone >28 mg | 0 (0) | 0 (0) |
| Dexamethasone 28 mg | 8 (0) | 3773 (54) |
| Dexamethasone 24 mg | 794 (6) | 298 (4) |
| Dexamethasone 16 mg | 10 (0) | 43 (1) |
| Dexamethasone 4-8 mg | 7562 (58) | 92 (1) |
| None | 4613 (36) | 774 (11) |
| **Discharged within 24 hours** | 2370 (18) | 2896 (41) |

Number of patients (% of all patients), except for Age, which is mean (SD). NSAID = non-steroidal anti-inflammatory drug, mg = milligram.

### Table S3: Subgroup analyses by type of surgery.

| **Outcomes** | **Control group (THA: n=** **10,576)**  **(KA: n=** **2,434)** | **Intervention group (THA: n=12,360)**  **(KA: n=21,947)** | **Estimated difference (minimal model)** | **Estimated difference (fully adjusted model)** |
| --- | --- | --- | --- | --- |
| **Primary outcome:**  **0-to-24-hour opioid consumption (mg IMEs)** |  |  |  |  |
| THA | 21.4 (22.9) | 20.3 (21.8) | -3.4 (95% CI -2.1;-4.7) | -1.0 (95% CI -0.1;-2.0) |
| KA | 25 (18.8) | 21 (20.8) | -0.7 (95% CI 0.5;-1.9) | -1.1 (95% CI -0.2;-2.0) |
| **Worst pain score within 0 to 24 hours (0-10 NRS)** |  |  |  |  |
| THA | 4.9 (2.4) | 4.8 (2.4) | -0.4 (99% CI -0.2;-0.6) | -0.3 (99% CI -0.1;-0.4) |
| KA | 3.9 (2.3) | 4.7 (2.4) | -0.5 (99% CI -0.3;-0.7) | -0.5 (99% CI -0.4;-0.7) |
| **Patients experiencing opioid-related adverse events (%)** |  |  |  |  |
| THA | 18.4 | 18.7 | -0.5 (99% CI 1.7;-2.7) | -1.0 (99% CI 1.1;-3.1) |
| KA | 7.9 | 17.5 | 6.5 (99% CI 9.2;3.8) | 5.6 (99% CI 8.1;3.1) |
| **Length of stay in the hospital (hours)** |  |  |  |  |
| THA | 36.4 (31.2) | 41.9 (59) | 0.7 (99% CI 4.5;-3) | 1.8 (99% CI 5.5;-1.9) |
| KA | 34.3 (17.4) | 37.6 (39.3) | -2.4 (99% CI 0.5;-5.3) | -2.5 (99% CI 0.4;-5.3) |
| **Days alive and out of hospital within 30 days** |  |  |  |  |
| THA | 28.1 (2.5) | 27.7 (3.4) | 0.0 (99% CI 0.3;-0.2) | 0.0 (99% CI 0.2;-0.3) |
| KA | 28.3 (1.6) | 28 (2.6) | 0.0 (99% CI 0.2;-0.2) | 0.0 (99% CI 0.2;-0.2) |

Baseline values are mean (standard deviation) or incidence in the control group. Estimates are adjusted mean differences in the intention-to-treat population. The minimal model is adjusted for time period and centre (random effect). The fully adjusted model is adjusted for age, sex, time period, type of anaesthesia, number of other multimodal analgesics, local infiltration analgesia, chronic opioid use, psychiatric diagnoses, and centre (random effect).

IME = Intravenous morphine equivalents, THA = Total hip arthroplasty, KA = Knee arthroplasty, CI = Confidence interval, NRS = Numerical rating scale score

### Table S4: Summary of findings in sensitivity analyses

| **Exclusion of participants with length of hospital stay under 24 hours** | | | | |
| --- | --- | --- | --- | --- |
| **Outcome** | **Control group (n=10,629)** | **Intervention group (n=25,379)** | **Estimated difference (minimal model)** | **Estimated difference (fully adjusted model)** |
| 0-to-24-hour opioid consumption (mg IMEs) | 22.8 (22.6) | 22.7 (21.9) | -3.9 (99% CI -3.1;-4.7) | -3.5 (95% CI -2.9;-4) |
| **Use of** **acetaminophen, NSAID and/or gabapentin as individual binary co-variables (instead of count)** | | | | |
| **Outcome** | **Control group (n=13,010)** | **Intervention group (n=34,307)** | **Estimated difference (minimal model)** | **Estimated difference (fully adjusted model)** |
| 0-to-24-hour opioid consumption (mg IMEs) | 22.1 (22.3) | 20.7 (21.2) | - | -3.5 (95% CI -3;-4) |
| **Complete case analysis of opioid-related adverse events within 0 to 24 hours** | | | | |
|  | **Control group (n=13,007)** | **Intervention group (n=34,265)** | **Estimated difference (minimal model)** | **Estimated difference (fully adjusted model)** |
| Patients experiencing opioid-related adverse events (%) | 16.4 | 17.9 | -1.1 (95% CI 0.2;-2.4) | -0.4 (95% CI 0.8;-1.7) |
| **Complete case analysis of worst pain score within 0 to 24 hours (0-10 NRS)** | | | | |
|  | **Control group (n=9,395)** | **Intervention group (n=25,015)** | **Estimated difference (minimal model)** | **Estimated difference (fully adjusted model)** |
| Worst pain score within 0 to 24 hours (0-10 NRS) | 5.3 (2.3) | 5.1 (2.4) | -0.9 (99% CI -0.8;-1) | -0.8 (99% CI -0.7;-0.9) |

Values are mean (standard deviation) or incidence. Estimates are adjusted mean differences. The minimal model is adjusted for time period, type of surgery, and centre (random effect). The fully adjusted model is adjusted for age, sex, time period, type of surgery, type of anaesthesia, number of other multimodal analgesics, local infiltration analgesia, chronic opioid use, psychiatric diagnoses, and centre (random effect).

IME = Intravenous morphine equivalents, CI = Confidence interval, NRS = Numerical rating scale score

### Figure S1: Run chart of the use of high-dose glucocorticoids relative to the defined time of implementation, at centres that implemented high-dose glucocorticoids during the study period.

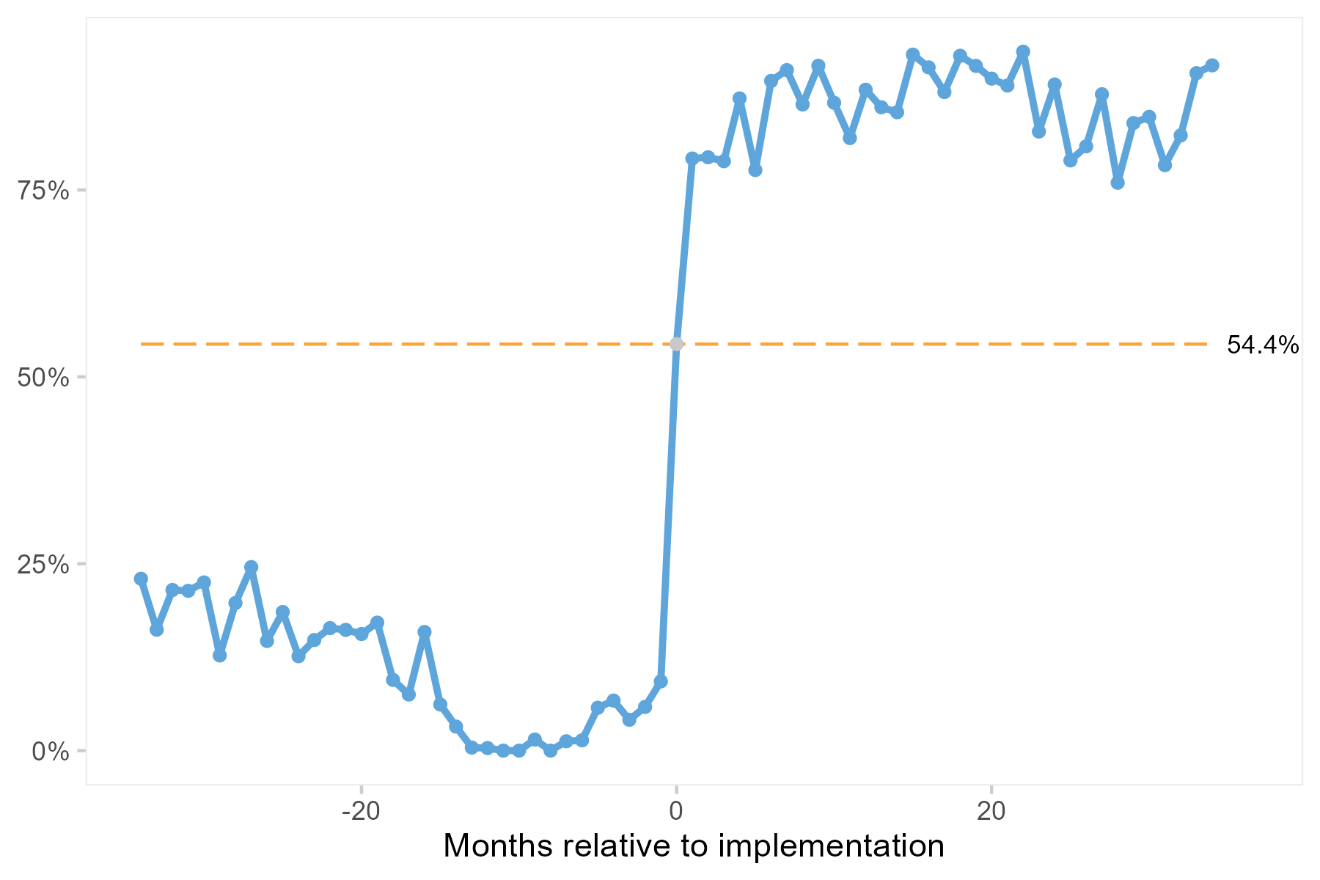

The chart includes the period from 34 months before to 34 months after the date of implementation, because the longest step was 34 months. The orange dashed line indicate the mean. The use of high-dose glucocorticoids that is seen from 34 to approximately 15 months before implementation stems from the inclusion of patients in the DEX2-TKA and RECIPE randomised trials, which randomised patients to 24 mg intravenous dexamethasone. Data points closer to time 0 are more important in the analyses, since the steps vary in size from 1 to 34 months.

Figure S2a-j: Statistical process control charts of co-variables relative to the defined time of implementation, at centres that implemented high-dose glucocorticoids during the study period. The charts include the period from 34 months before to 34 months after the date of implementation, because the longest step was 34 months. Orange dashed mean lines indicate ‘unusually’ few crossings over the mean or ‘unusually’ many consecutive data points on the same side of the mean. Orange points outside the grey 3-sigma control limits. Both indicate non-random variation.

2a: **Participants age**

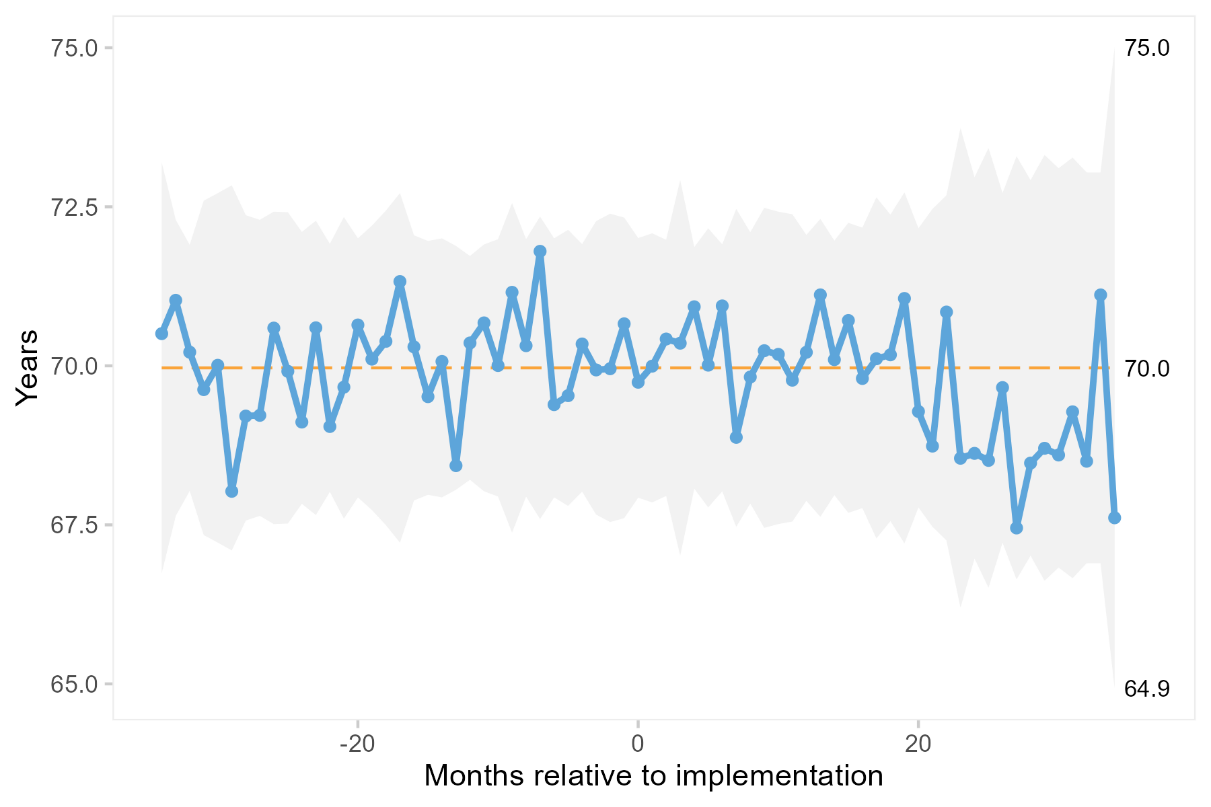

2b: **Proportion of female participants**

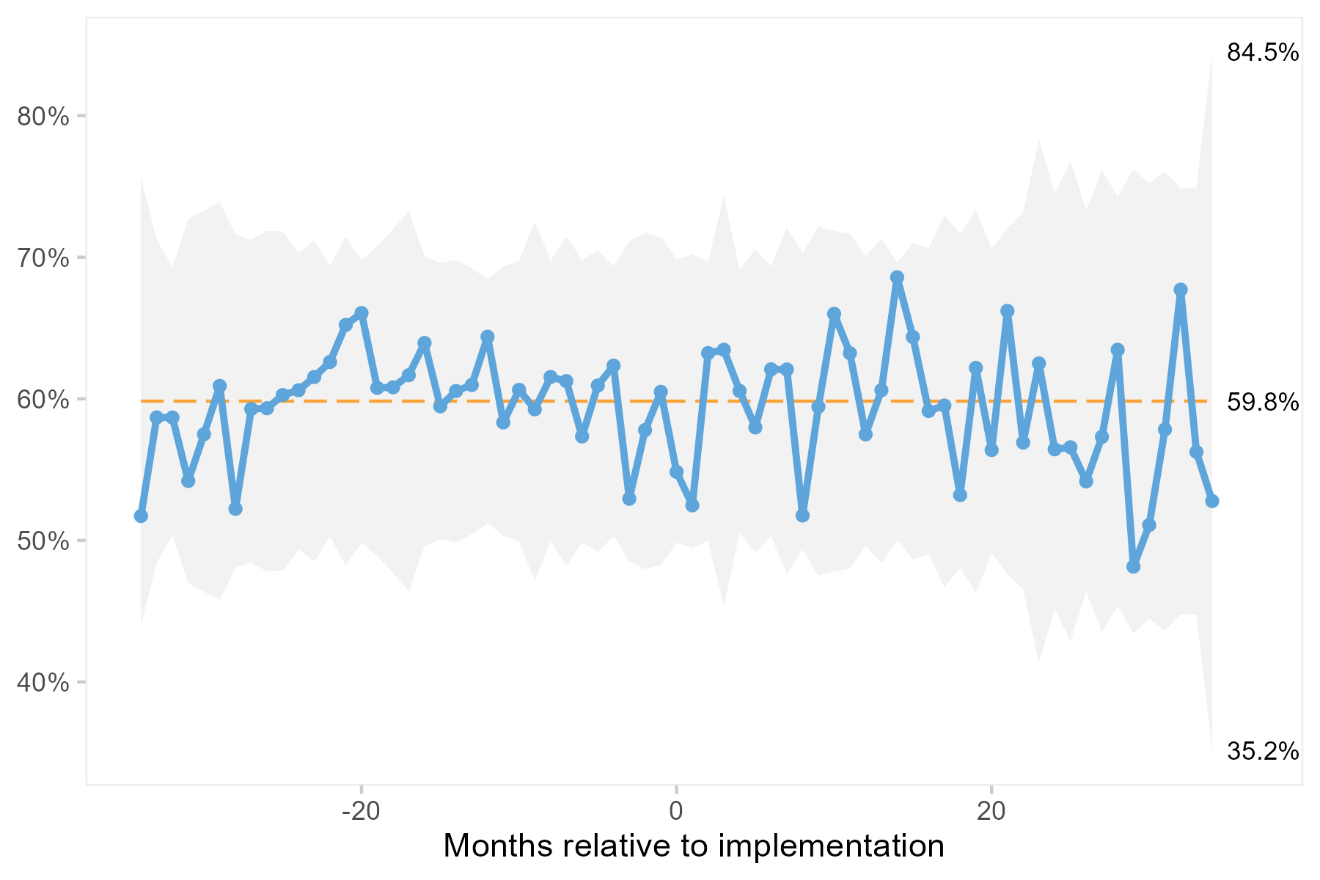

2c: **Proportion of participants with chronic preoperative opioid use**

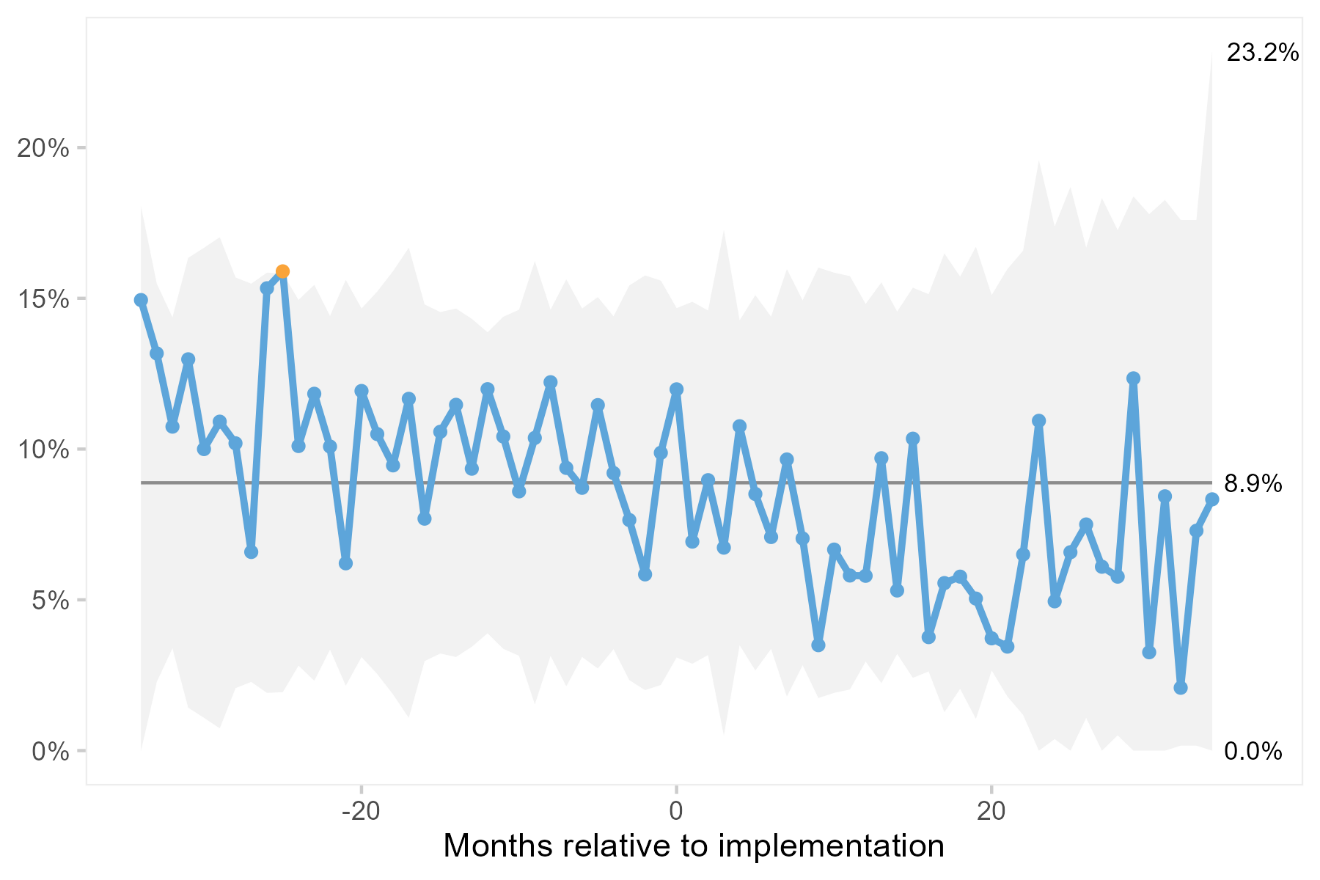

2d: **Proportion of participants with a psychiatric diagnosis**

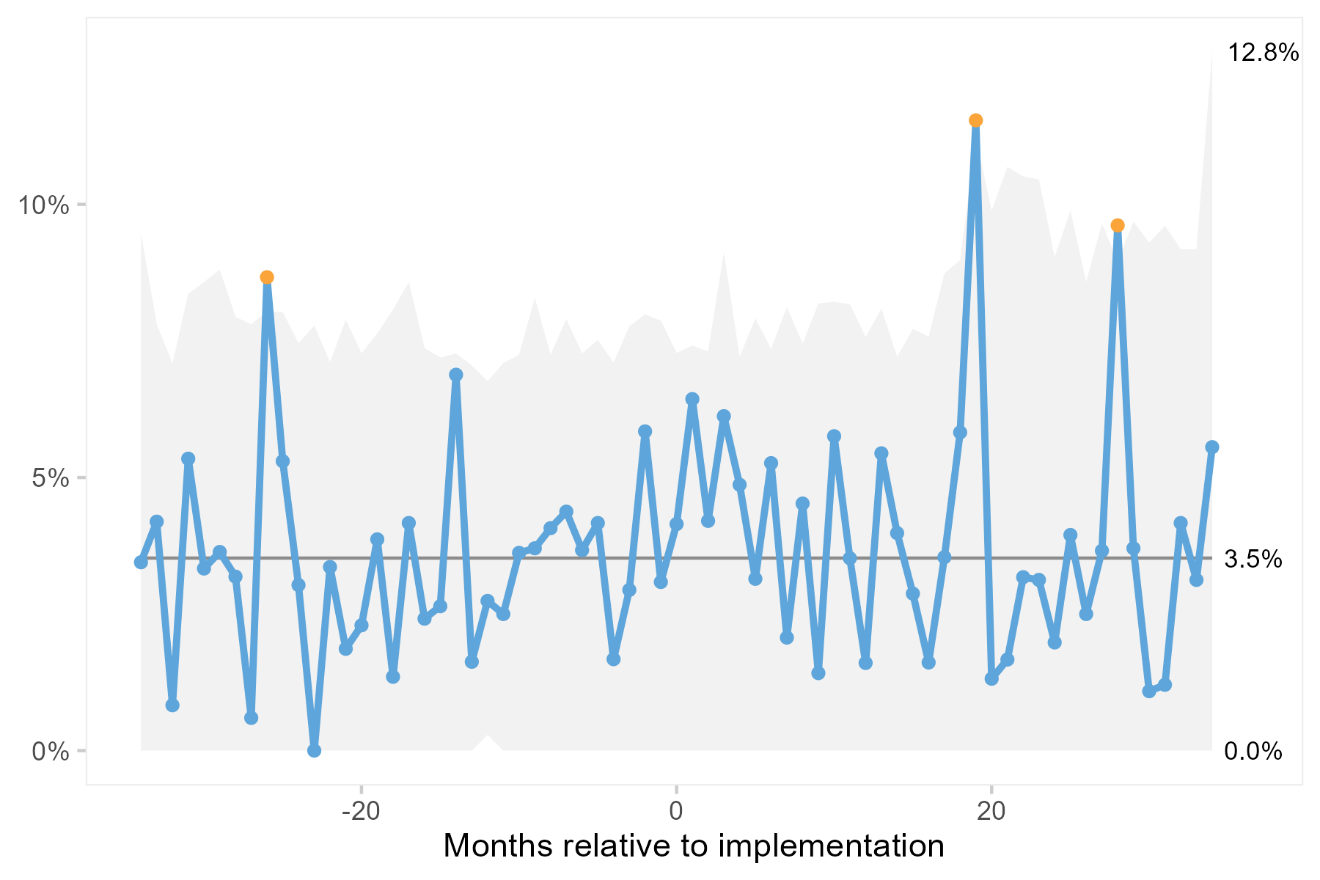

2e: **Proportion of participants treated with spinal anaesthesia**

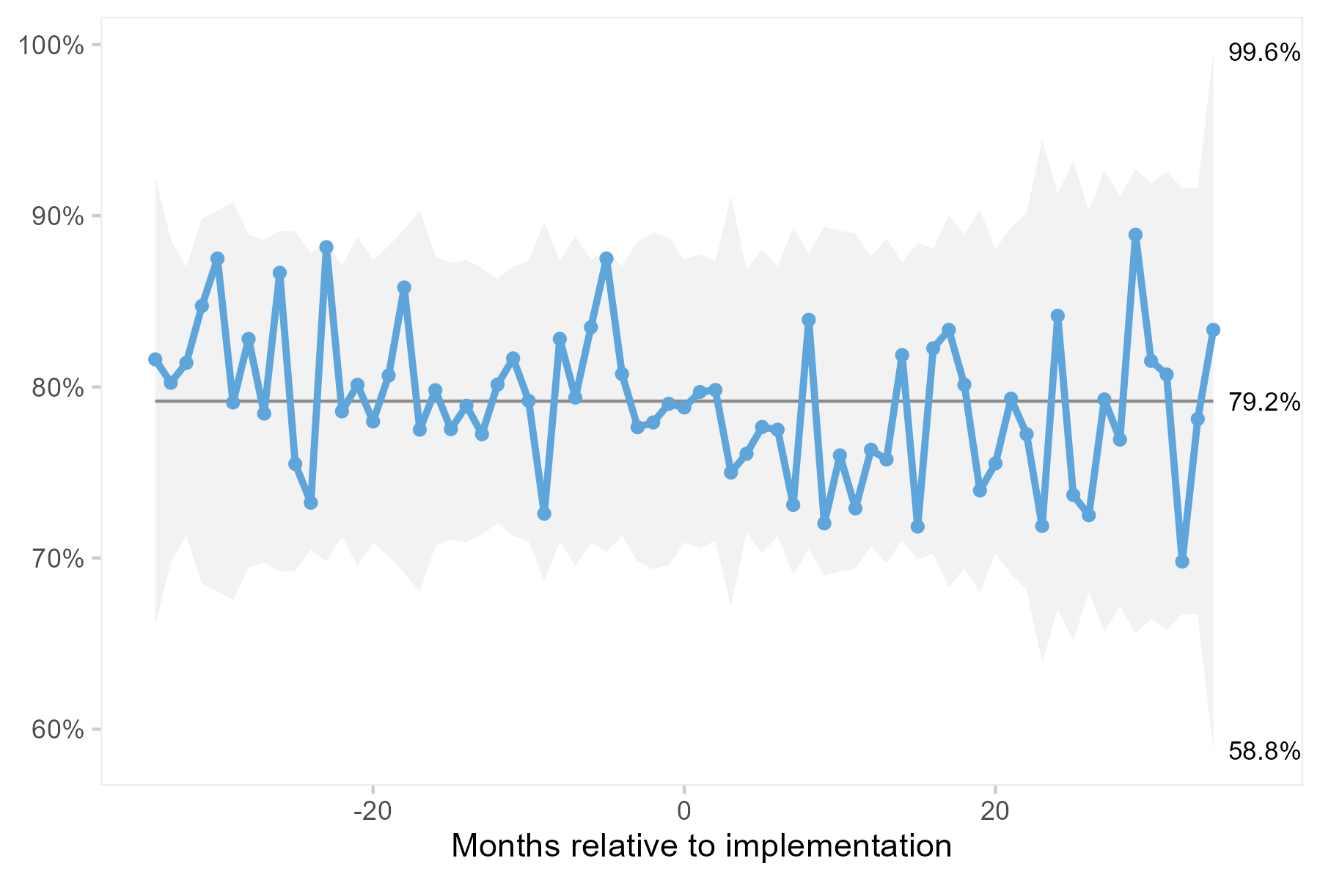

2f: **Proportion of participants treated with general anaesthesia and a nerve block**

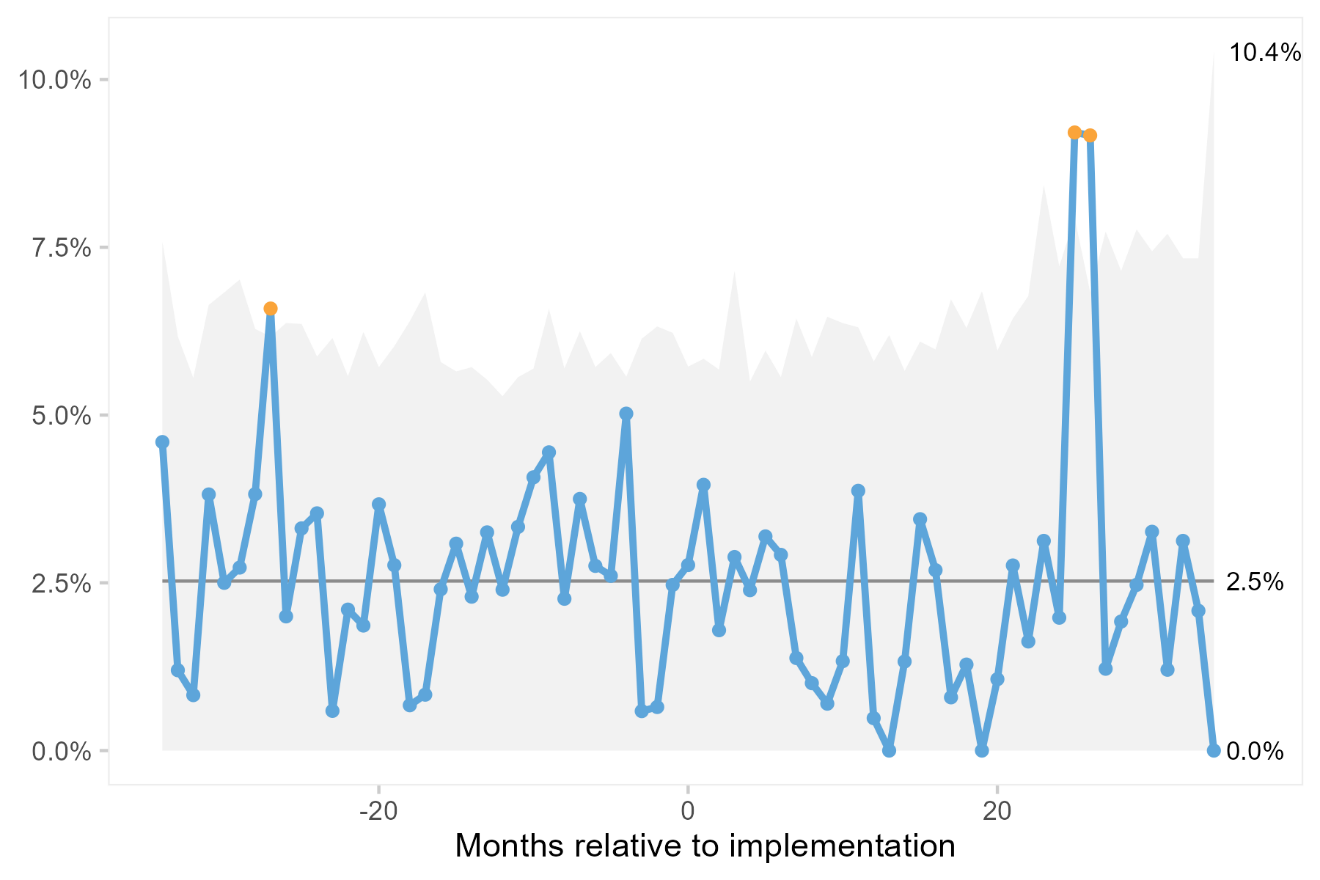

2g: **Proportion of participants treated local infiltration analgesia**

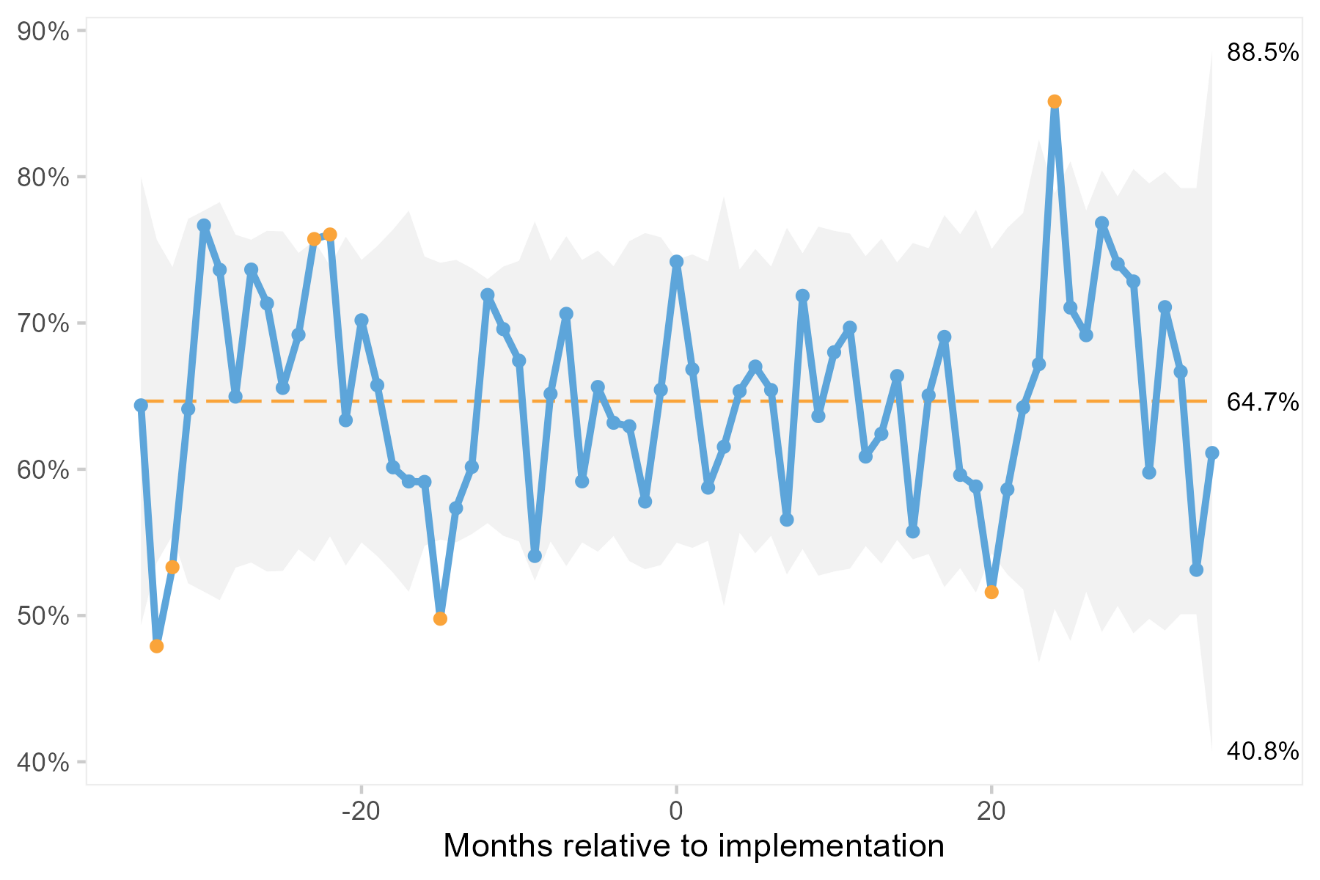

2h: **Proportion of participants treated with acetaminophen**

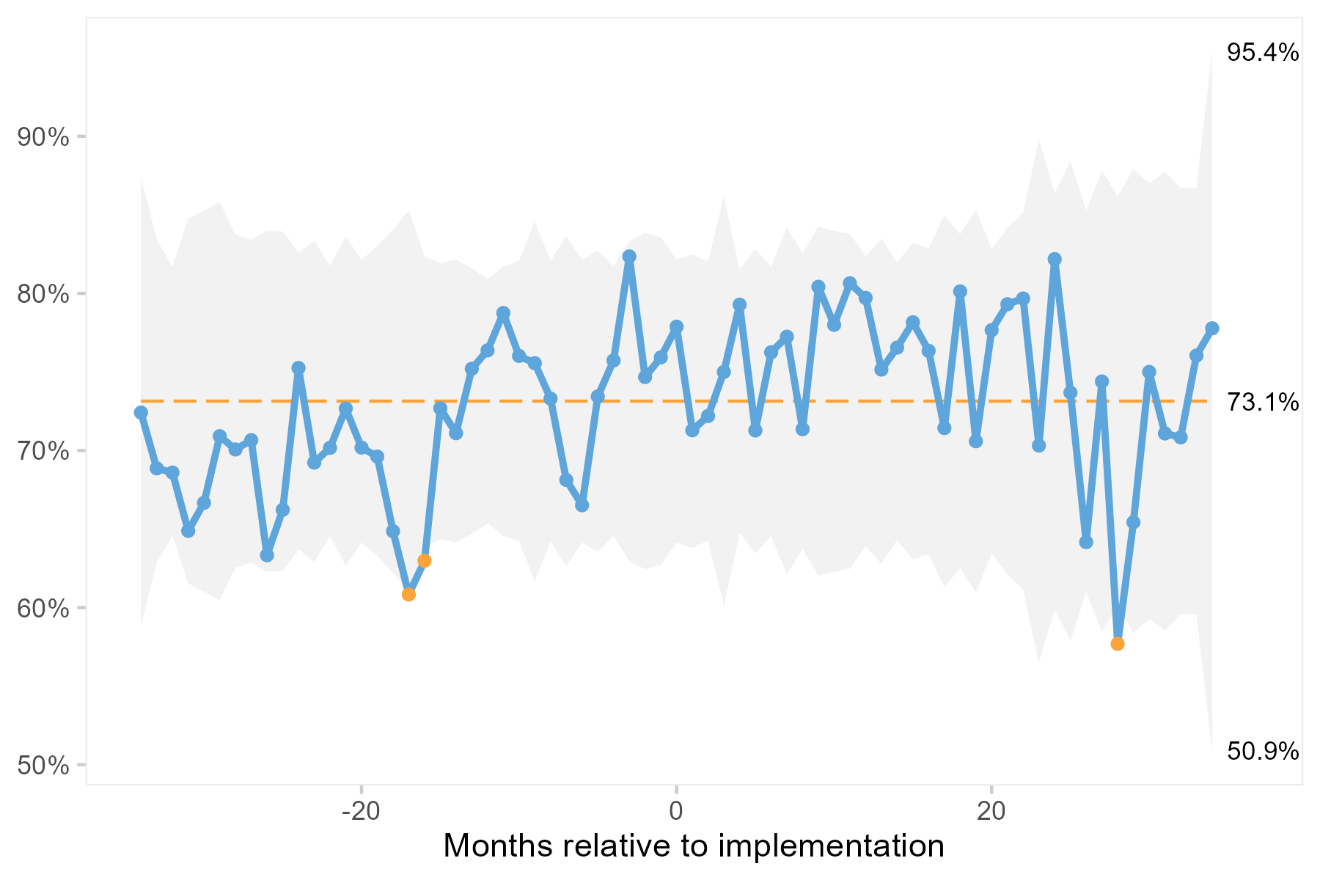

2i: **Proportion of participants treated with a non-steroidal anti-inflammatory drug (NSAID)**

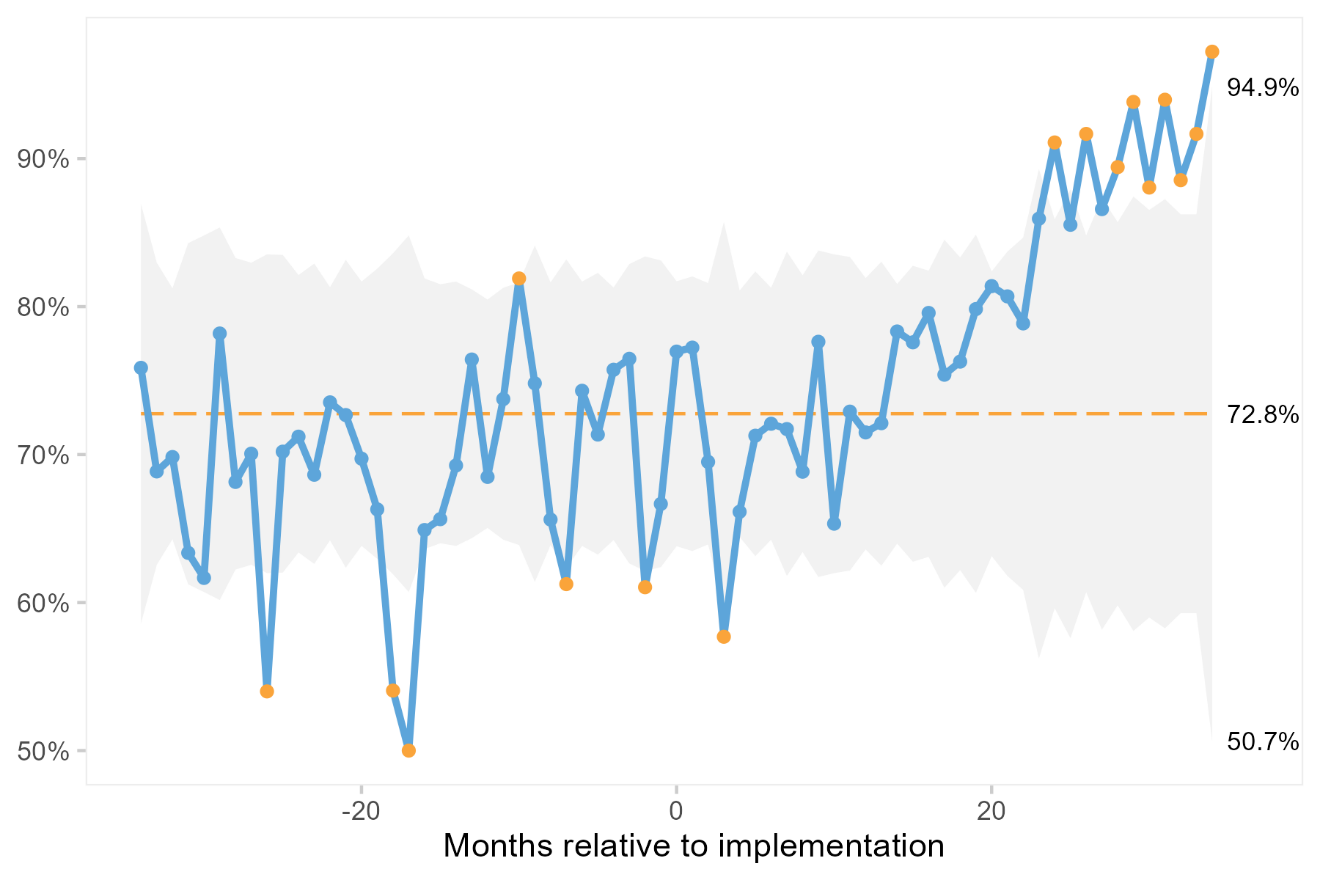

2j: **Proportion of participants treated with a gabapentinoid**

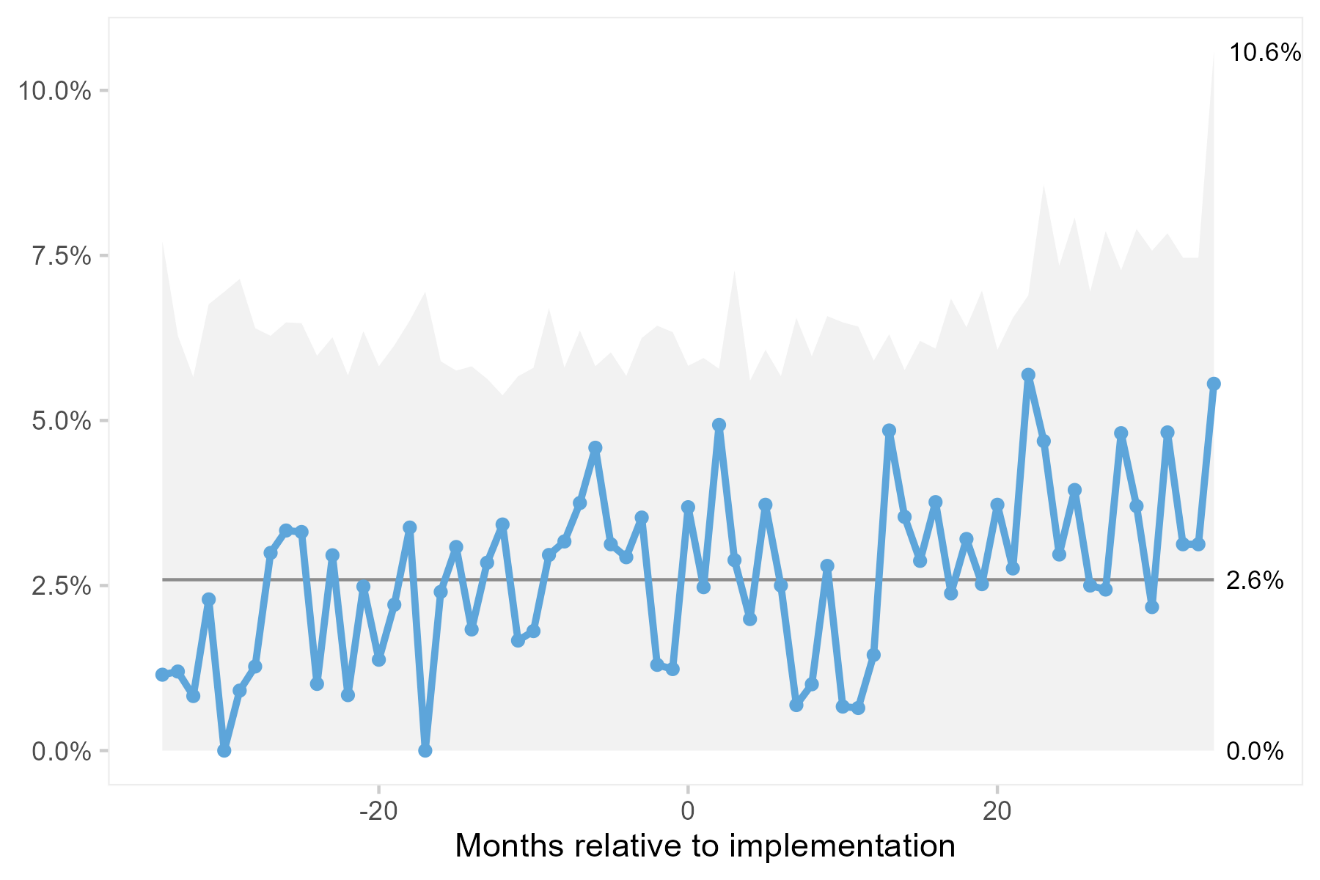

Figure S3 a-e: Statistical process control charts of outcome measures relative to the defined time of implementation, at centres that implemented high-dose glucocorticoids during the study period. Orange dashed mean lines indicate ‘unusually’ few crossings over the mean or ‘unusually’ many consecutive data points on the same side of the mean. Orange points outside the grey 3-sigma control limits. Both indicate non-random variation.

3a: O**pioid consumption in the first 24 hours after surgery, expressed in intravenous morphine equivalent doses (IME)**

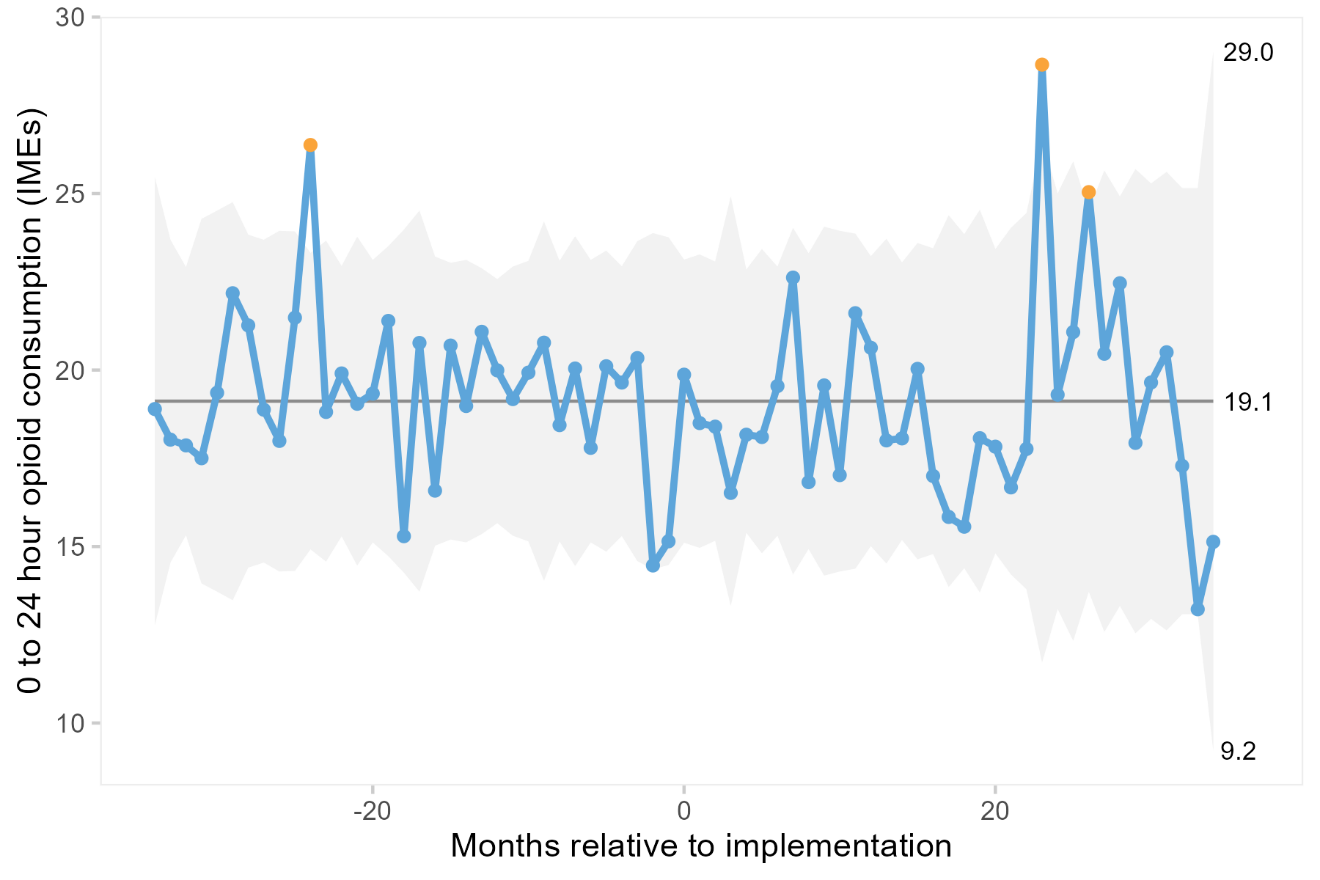

3b: **Worst postoperative pain intensity during 0-24 hours after surgery, measured with the 0-10 numerical rating scale (NRS)**

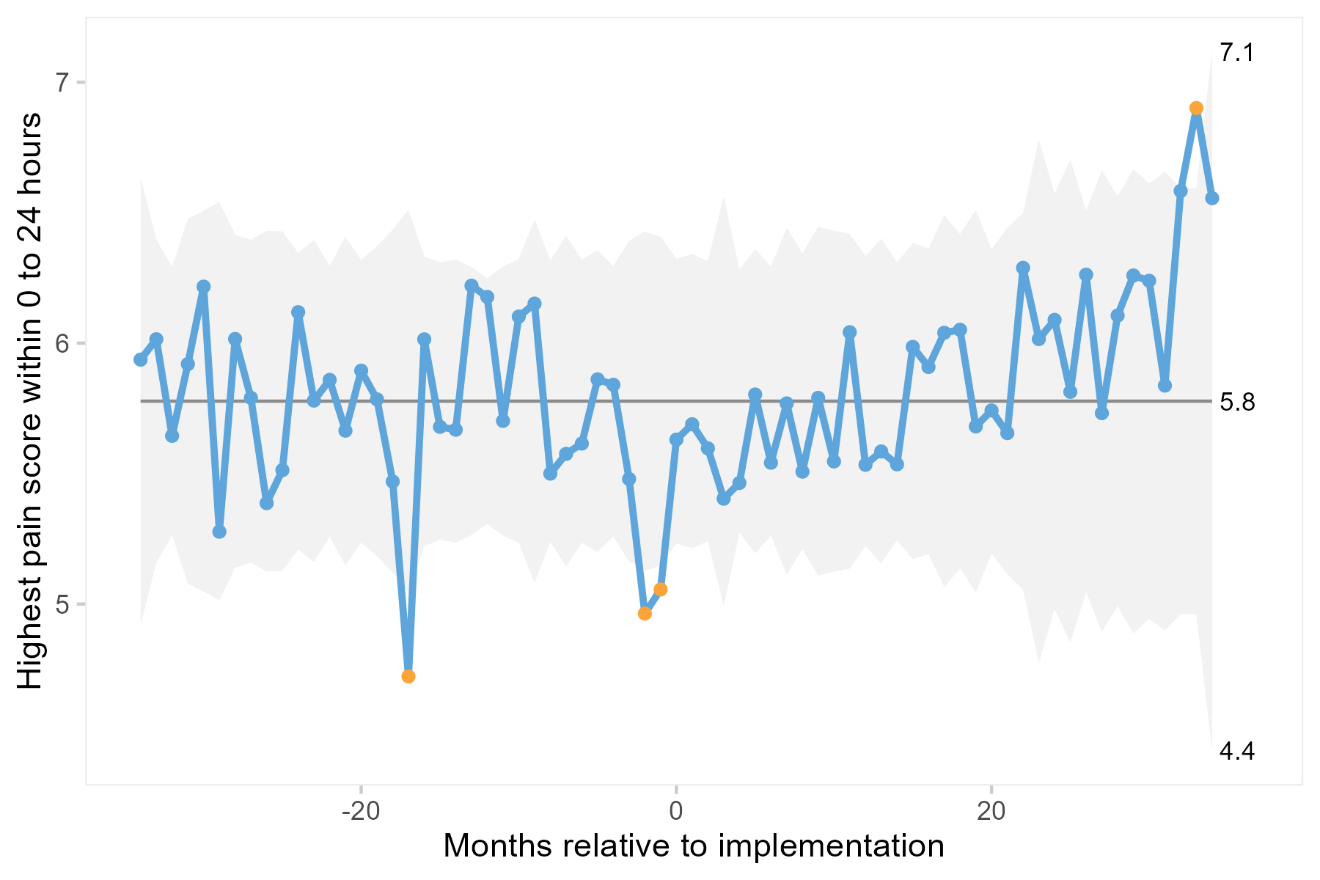

3c: **Proportion of patients with at least one opioid-related adverse event (ORADE) during 0-24 hours after surgery**

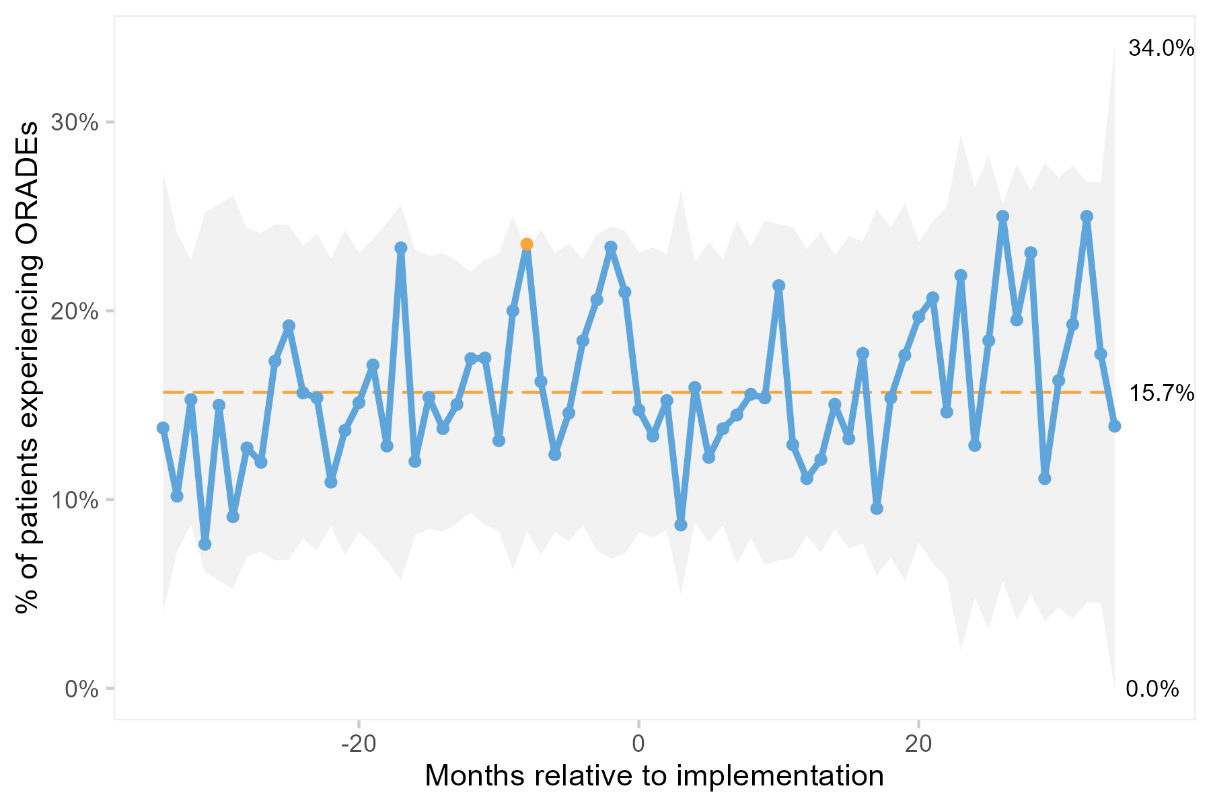

3d: **Length of hospital stay, measured in hours from end of surgery to hospital discharge**

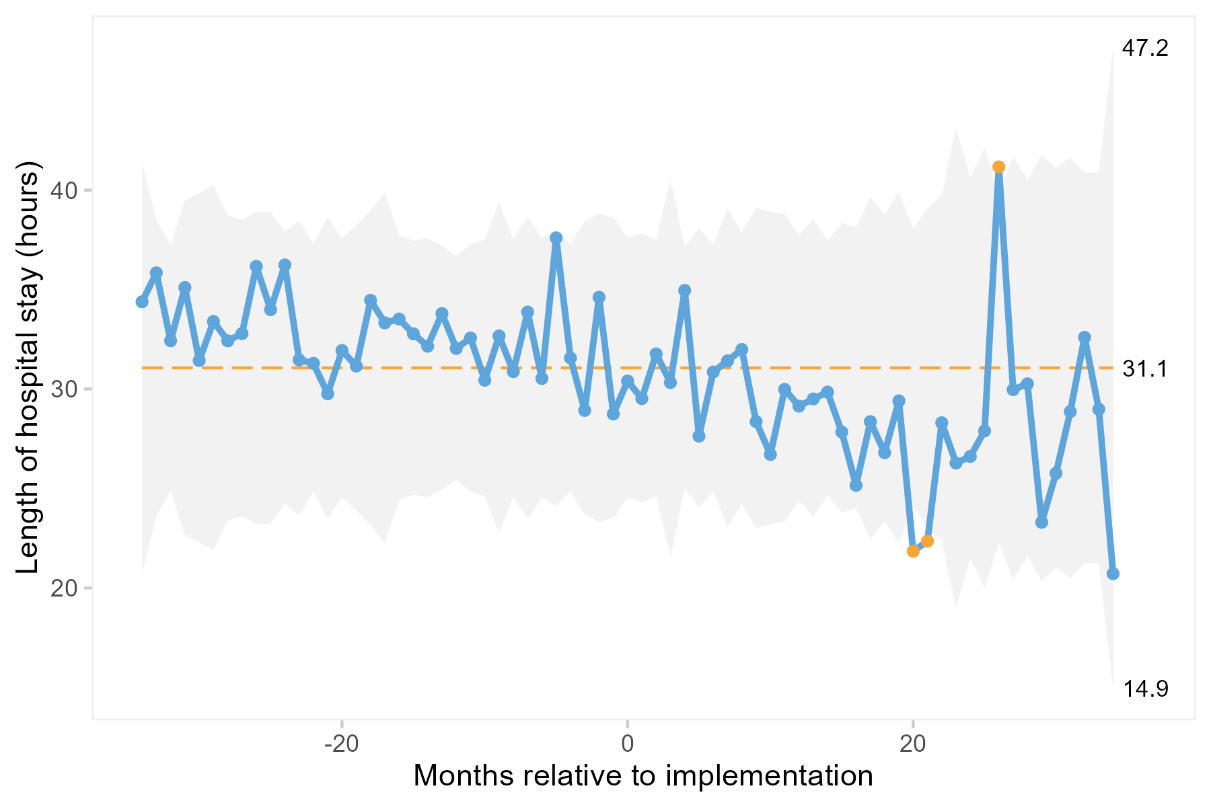

3e: **Days alive and out of hospital at 30 days**

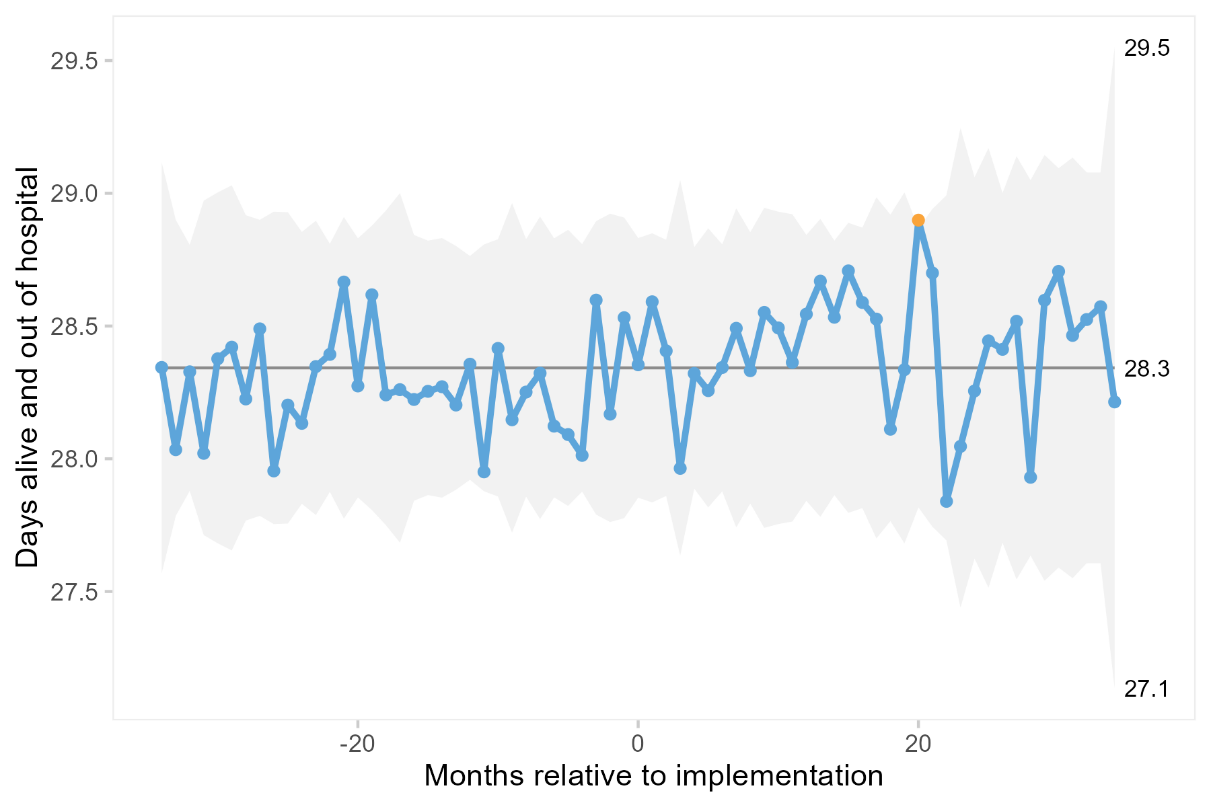
